# One-year within-trial and lifetime-horizon modeled health economic evaluation of the risk-stratified Prediabetes Lifestyle Intervention Study (PLIS) for prediabetes remission in Germany

**DOI:** 10.64898/2026.05.22.26353768

**Authors:** Damon Mohebbi, Markus Vomhof, Joseph Montalbo, Ann Kathrin Winkels, Veronika Gontscharuk, Nadja Chernyak, Charalabos-M. Dintsios, Nadja Kairies-Schwarz, Renée Stark, Karl M.F. Emmert-Fees, Min Fan, Renate Schick, Annette Schürmann, Stefan Bornstein, Martin Heni, Norbert Stefan, Reiner Jumpertz von Schwartzenberg, Matthias Blüher, Andreas Lechner, Julia Clavel, Stefan Kopf, Julia Szendrödi, Michael Roden, Robert Wagner, Andreas Fritsche, Andreas L. Birkenfeld, Andrea Icks

## Abstract

**Background:** Lifestyle interventions can increase the probability of remission of prediabetes to normal glucose tolerance, but their economic value remains unclear. We assessed the within-trial and lifetime-horizon modeled cost-effectiveness of intensive and conventional lifestyle interventions in risk-stratified participants with prediabetes.

**Methods:** A health economic evaluation was conducted alongside the 12-month multicenter PLIS trial (n=1,105). High-risk participants were randomized to intensive (HR-INT) or conventional (HR-CONV); low-risk participants to conventional lifestyle intervention (LR-CONV) or control (only short single consultation; LR-CTRL) with risk stratification based on insulin secretion, insulin sensitivity, and liver fat content. Within-trial analyses estimated incremental costs per additional remission to normoglycemia and per quality-adjusted life year (QALY). Lifetime cost-effectiveness was modelled using a four-state Markov Model.

**Findings:** At 12 months, HR-INT and LR-CONV increased remission compared with their respective comparators. The incremental cost per additional remission was €7,081 (95% CI: dominated-47,277) for HR-INT and €4,278 (1,312–11,793) for LR-CONV from a health insurance perspective. A willingness-to-pay of €22,000 (HR-INT) and €7,500 (LR-CONV) per additional remission corresponded to 90% probability of cost-effectiveness. Neither intervention was cost-effective in terms of QALYs gained within the 12-months period. Lifetime modelling suggested that both HR-INT and LR-CONV are not only cost-effective, but also cost-saving, relative to HR-CONV and LR-CTRL, respectively. Also in the probabilistic sensitivity analysis, most simulations indicated dominance (71.7% for HR and 88% for LR).

**Interpretation:** Based on short-term economic evaluation, the interventions assessed were cost-effective regarding additional participants with remission, not for incremental QALYs gained. Lifetime modelling suggests cost savings for both risk groups. Targeting populations with lifestyle interventions to achieve prediabetes remission seems to generate good value for money in the long term.

*Trial registration number:* NCT01947595

**Research in context:** *What is already known about this subject?:*

- Lifestyle modification is pivotal for type 2 diabetes prevention in individuals with prediabetes; improvements are needed to overcome nonresponse to preventive interventions.
- The 12-month Prediabetes Lifestyle Intervention Study (PLIS) was the first multicenter study, which involved eight study sites in university hospitals in Germany, where investigators prospectively tested different intensities of lifestyle intervention in a risk-stratified manner.
- PLIS achieved higher remission rates of prediabetes with an intensive lifestyle intervention (16 counseling sessions) in high-risk participants and a conventional lifestyle intervention (8 counselling sessions) in low-risk participants, relative to a conventional (8 counselling sessions) and control lifestyle intervention (single short counselling session), respectively.
- As risk-stratified diabetes prevention has not been implemented so far and more intense lifestyle interventions are usually associated with higher healthcare utilization, it remains unclear whether these interventions are cost-effective.

*What is the key question?:*

- Are intensive and conventional lifestyle interventions of the Prediabetes Lifestyle Intervention Study cost-effective after 12 months and over the lifetime horizon?

*What are the new findings?:*

- Intensive lifestyle intervention in high-risk participants and the conventional lifestyle intervention in low-risk participants are both likely to be cost-effective at a willingness-to-pay threshold between €10,000 and €20,000 per additional person with prediabetes remission.
- Neither intensive lifestyle intervention nor conventional lifestyle intervention are cost-effective in terms of quality adjusted life years gained in the 12-month trial period; however, this was not unexpected, given the limited scope for short-term improvement in health-related quality of life in a largely asymptomatic population with high baseline utility values.
- Simulation modeling with a lifetime horizon thereby shows that intensive lifestyle intervention in high-risk participants and conventional lifestyle intervention in low-risk participants may be not only cost-effective but also cost-saving.

*How might this impact on clinical practice in the foreseeable future?:*

- Individualized, risk phenotype-based lifestyle intervention in prediabetes may be cost-effective regarding remission to normal glucose regulation in the short- and regarding quality-adjusted life years gained the long-term.
- When evaluating lifestyle-based type 2 diabetes prevention, a long-term perspective is essential, as the intervention may translate into clinically meaningful and economically relevant benefits only over time through delayed diabetes onset, fewer diabetes-related complications, and reduced healthcare utilization.

## Introduction

Type 2 diabetes mellitus (T2DM) is associated with multiple complications, high mortality rates, and high healthcare costs (1) The development of T2DM is commonly understood as part of a continuum of dysglycemia, progressing from normal glucose tolerance to prediabetes and, ultimately, overt T2DM. Prediabetes refers to elevated blood glucose concentrations that are below the defined thresholds for diabetes diagnosis according to the criteria of the American Diabetes Association (ADA) (2). Studies indicate that the progression of prediabetes to diabetes is likely associated with a family history of diabetes, male sex, and high systolic blood pressure (3,4).

Identifying people with prediabetes and preventing or delaying the progression to diabetes is important from the healthcare and societal perspective. Some prevention strategies involve pharmacological or surgical interventions, lifestyle interventions (LIs) focus on promoting physical activity, a healthy diet, and body weight reduction. Studies have shown that lifestyle interventions effectively reduce the risk of developing diabetes in people with prediabetes (5) and tend to be cost-effective in health economic evaluations (6,7). Recently, glycemic remission has been advocated as a goal for preventing T2DM in people with prediabetes (8).

Intensive lifestyle interventions may be particularly beneficial in participants with prediabetes at high risk of developing diabetes. In this respect, individuals with a high-risk phenotype of prediabetes (based on low disposition index and/or low insulin sensitivity with nonalcoholic fatty liver disease) responded less favorably to conventional lifestyle interventions since their blood sugar levels only improved slightly for the same amount of weight loss (9).

Given that intensive lifestyle interventions are usually associated with higher healthcare utilization and costs than conventional lifestyle interventions, it remains unclear whether these interventions are cost-effective in participants with only a high-risk prediabetes phenotype. Up to now, only few studies have come close to addressing this question: Previous simulation models in the UK have shown lifestyle interventions targeting high-risk individuals without or with prediabetes are likely to be cost-effective, but this can improve (10) or deteriorate (11) depending on intervention intensity. An economic evaluation of a Dutch lifestyle intervention in the primary care setting showed cost-effectiveness compared to usual health care (12).

The Prediabetes Lifestyle Intervention Study (PLIS), a randomized, controlled, multicenter intervention study over 12 months, was the first to prospectively test different intensities of lifestyle intervention in a risk-stratified population with prediabetes, published in 2021 (13). In a joint analysis from PLIS and the US Diabetes Prevention Program (DPP) the concept of “prediabetes remission” was established (14). Moreover, prediabetes remission was associated with a delay of T2DM by more than 6 years in DPP as well as reduced CV outcomes in DPP and the DaQing Diabetes Prevention Study (15,16).

Achieving remission from prediabetes may therefore have long-term effects on health and cost outcomes, which within-trial economic evaluations based on short-term follow-up periods cannot adequately capture. Because individuals with prediabetes are often relatively healthy and typically do not yet experience the costly and clinically overt complications associated with T2DM, substantial effects on costs and health outcomes are generally not expected over short follow-up periods. To project the long-term impact of interventions, decision-analytic models are commonly used to extrapolate findings of clinical trials (17), such as the 12-month PLIS trial, by incorporating data from other sources including national life tables. Due to the absence of risk-stratified prediabetes remission models and the limited availability of German data, a pragmatic, simplified decision modeling approach is required to make a first step in addressing these gaps. Thus, in this study, we aim to assess both the within-trial and lifetime-horizon cost-effectiveness of two lifestyle interventions with different intensity in a risk-stratified population with prediabetes alongside the PLIS trial.

## Research Design and Methods

### The Prediabetes Lifestyle Intervention Study (PLIS)

The risk-stratified randomized-controlled trial (RCT) PLIS analyzed the effectiveness of (I) an intensive lifestyle intervention (HR-INT) compared to a conventional lifestyle intervention (HR-CONV) in individuals with a high-risk phenotype of prediabetes (low disposition index and/or low insulin sensitivity with nonalcoholic fatty liver disease) and the effectiveness of (II) a conventional lifestyle intervention (LR-CONV) compared to a control intervention (LR-CTRL) in participants with a low-risk phenotype of prediabetes (13). PLIS, which ran between 2012 and 2017, was approved by all local ethics committees of the participating institutions and involved eight study sites in university hospitals in Germany.

Out of 2561 participants assessed for eligibility (criteria: adults aged 18 to 75 years with clinically suspected prediabetes or at least 50 points in the German Diabetes Risk assessment battery (18)), 1149 participants were first risk-stratified according to insulin secretion, insulin sensitivity, and liver fat content (9) based on a 2-hour, 75 g OGTT and magnetic resonance spectroscopy and then randomly assigned to a study group. Over 12 months, high-risk participants received either HR-INT or HR-CONV, whereas low-risk participants received either LR-CONV or LR-CTRL (13). Out of 1149 risk-stratified participants, a total of 908 individuals (79.03%) with complete information (i.e., no missing values) from a 75-g oral glucose tolerance test (OGTT) were included in the analysis. The study population was composed of 707 high-risk participants (77.86%, HR-INT=356, HR-CONV=351) and 201 low-risk (22.14%, LR-CONV=100, LR-CTRL=101) participants.

HR-INT comprised 16 sessions of dietary and exercise counseling and recommended physical activity of at least 6 hours per week, HR-CONV and LR-CONV contained 8 sessions with a physical activity recommendation of at least 3 hours per week. Sessions lasted 30 to 60 minutes and were led by a lifestyle advisor (i.e., dietitian) with prior training. Participants in the LR-CTRL group only received a 30-minute consultation at the beginning of the study.

Data was collected at baseline, 6 months after baseline, and 12 months after baseline.

### Design of the within-trial health economic evaluation

A health economic evaluation was conducted alongside the PLIS trial, assessing the cost-effectiveness of the two lifestyle interventions with different intensity versus their respective comparators at the 12-month follow-up. This evaluation was reported according to the Consolidated Health Economic Evaluation Reported Standards Statement (CHEERS) (19). Cost-effectiveness was assessed through a cost-effectiveness analysis and a cost-utility analysis in terms of additional costs per participant with remission to normal glucose regulation and in terms of additional costs per quality-adjusted life year (QALY) gained, respectively. The analyses were carried out from the perspective of German Statutory Health Insurance (SHI) and a pragmatic societal perspective including participants’ out-of-pocket payments and time costs as well as indirect costs resulting from productivity losses. An impact inventory table (20) is provided in ESM Table 1. Because the PLIS trial lasted only a year, we did not discount costs and outcomes (21).

**Table 1.**
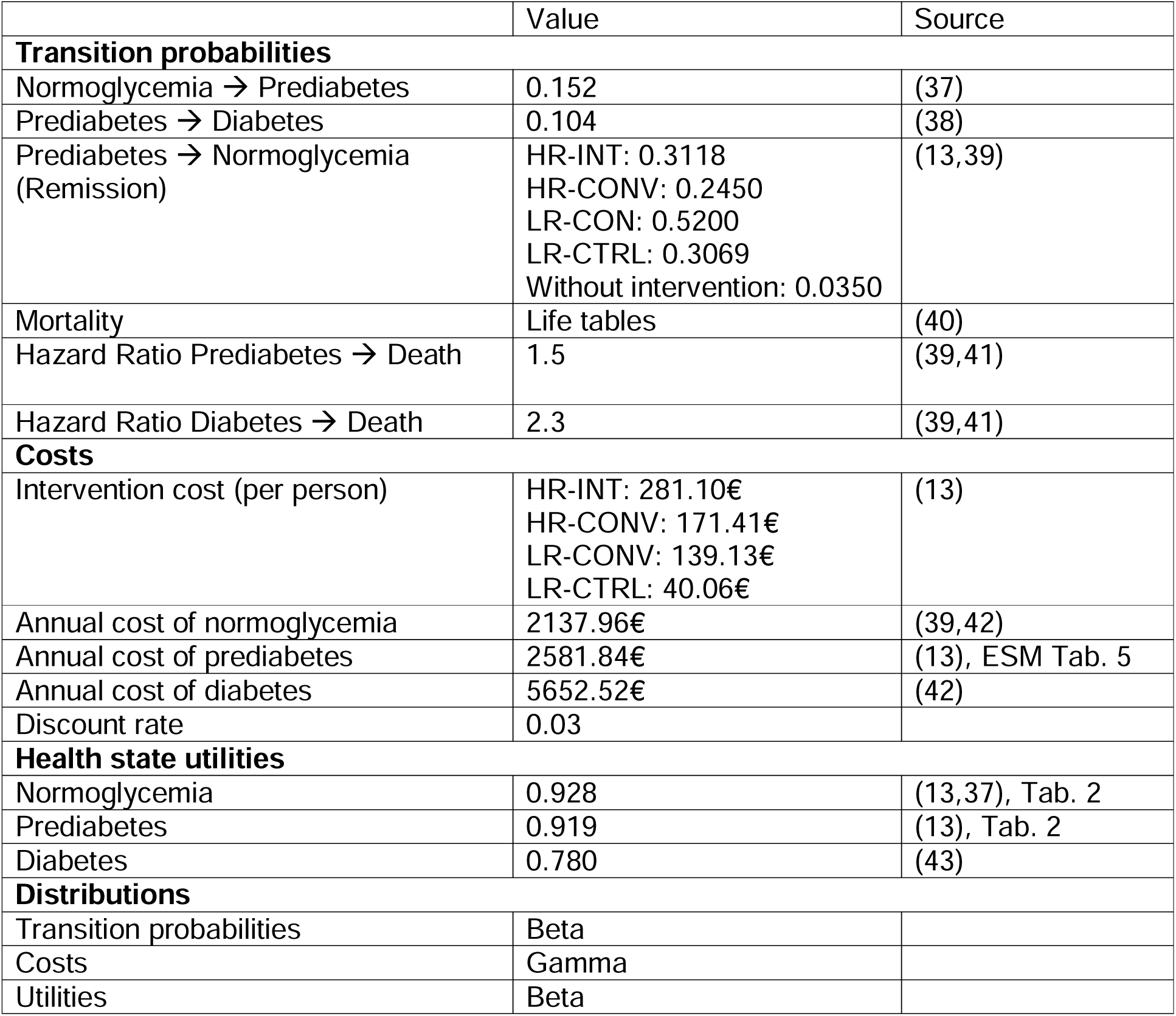
Input parameters for the lifetime model.

|  | Value | Source |
| --- | --- | --- |
| **Transition probabilities** |  |  |
| Normoglycemia → Prediabetes | 0.152 | (37) |
| Prediabetes → Diabetes | 0.104 | (38) |
| Prediabetes → Normoglycemia (Remission) | HR-INT: 0.3118<br>HR-CONV: 0.2450<br>LR-CON: 0.5200<br>LR-CTRL: 0.3069<br>Without intervention: 0.0350 | (13,39) |
| Mortality | Life tables | (40) |
| Hazard Ratio Prediabetes → Death | 1.5 | (39,41) |
| Hazard Ratio Diabetes → Death | 2.3 | (39,41) |
| **Costs** |  |  |
| Intervention cost (per person) | HR-INT: 281.10€<br>HR-CONV: 171.41€<br>LR-CONV: 139.13€<br>LR-CTRL: 40.06€ | (13) |
| Annual cost of normoglycemia | 2137.96€ | (39,42) |
| Annual cost of prediabetes | 2581.84€ | (13), ESM Tab. 5 |
| Annual cost of diabetes | 5652.52€ | (42) |
| Discount rate | 0.03 |  |
| **Health state utilities** |  |  |
| Normoglycemia | 0.928 | (13,37), Tab. 2 |
| Prediabetes | 0.919 | (13), Tab. 2 |
| Diabetes | 0.780 | (43) |
| **Distributions** |  |  |
| Transition probabilities | Beta |  |
| Costs | Gamma |  |
| Utilities | Beta |  |

The outcome measure of the cost-effectiveness analysis was related to prediabetes remission. We generated a binary outcome variable indicating whether a person’s blood glucose level had returned to normal at the 12-month follow-up (i.e., participant with remission to normal glucose regulation and, thus, a participant without prediabetes or diabetes) according to ADA classification (2).

We calculated utility weights by evaluating the self-assessed health states of participants resulting from the EQ-5D-3L and the EQ-5D-5L questionnaires by applying a German tariff (ESM Table 2) (22,23). To determine total QALYs over the 12-months study period, we calculated the area under the curve by applying linear interpolation between utility weights (24) measured at baseline, 6 months after baseline, and 12 months after baseline. Costs were calculated in Euros (€) for the reference year 2017 (the last the year the intervention took place) by multiplying quantities and prices from published sources and official German statistics. A breakdown of prices by type of costs and perspective is provided in ESM Table 3. Details on how we calculated costs are provided in ESM Table 4, and a calculation of intervention-related costs is provided in ESM Table 5.

**Table 2.**
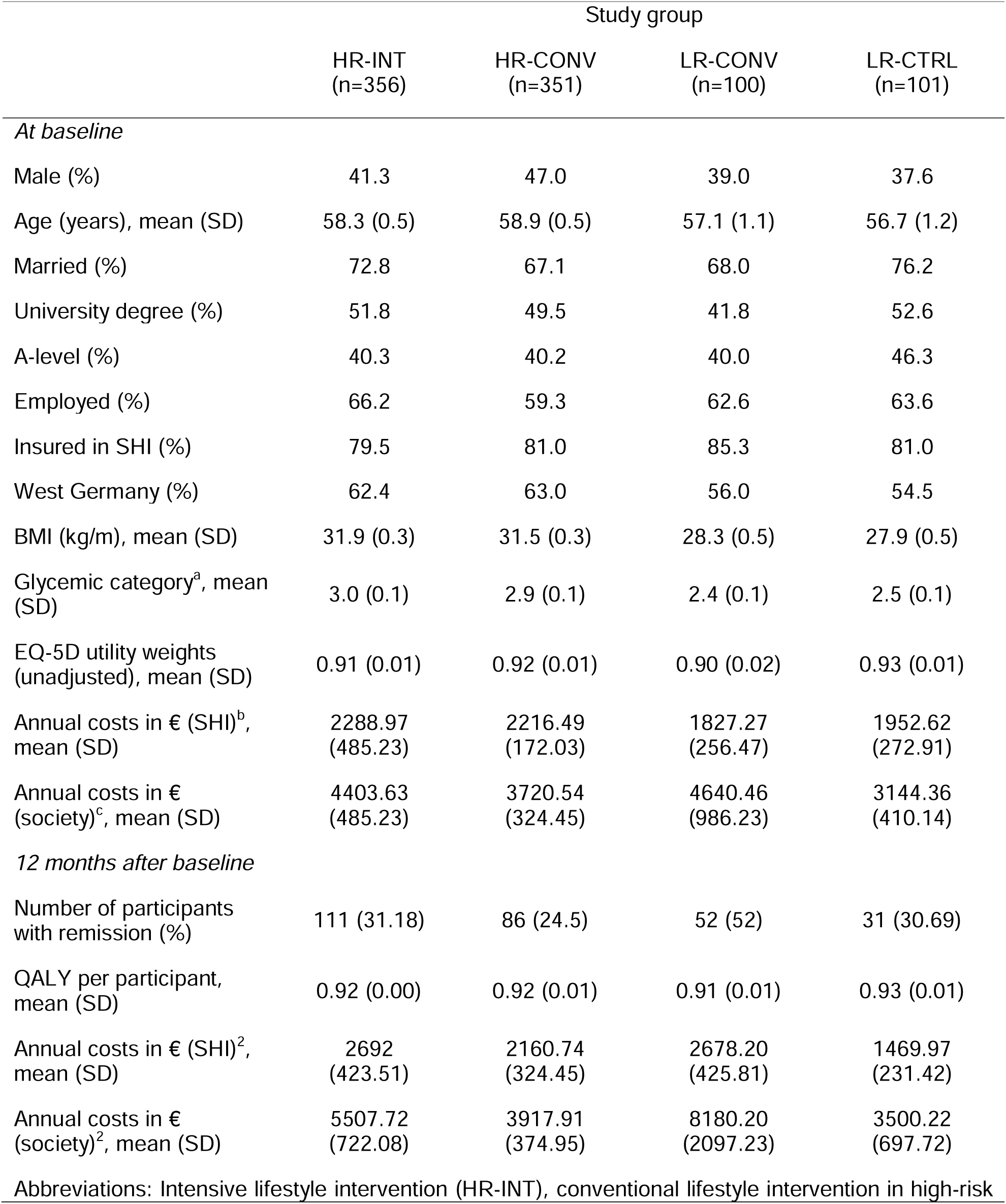
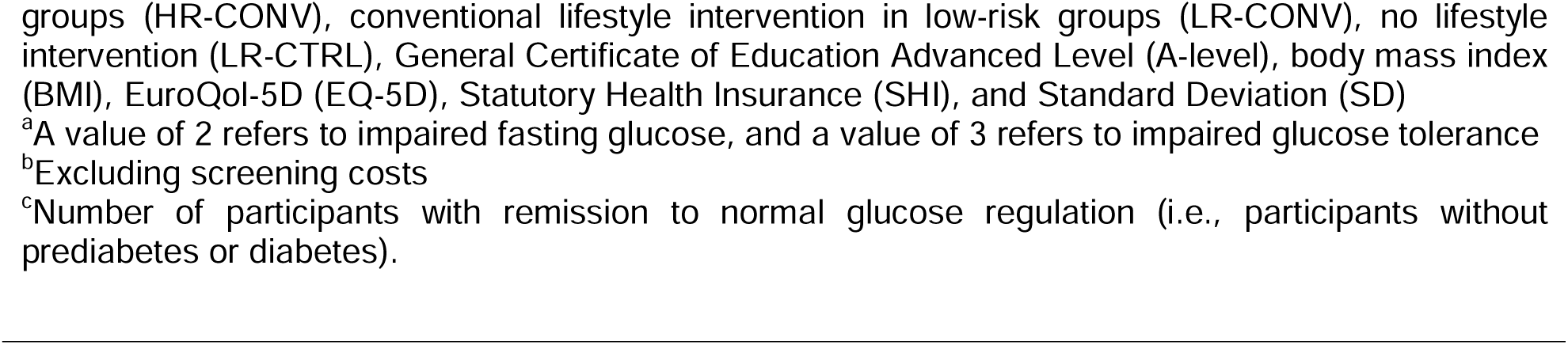
Description of the population at baseline and 12 months after baseline.

|  | Study group |  |  |  |
| --- | --- | --- | --- | --- |
|  | HR-INT<br>(n=356) | HR-CONV<br>(n=351) | LR-CONV<br>(n=100) | LR-CTRL<br>(n=101) |
| *At baseline* |  |  |  |  |
| Male (%) | 41.3 | 47.0 | 39.0 | 37.6 |
| Age (years), mean (SD) | 58.3 (0.5) | 58.9 (0.5) | 57.1 (1.1) | 56.7 (1.2) |
| Married (%) | 72.8 | 67.1 | 68.0 | 76.2 |
| University degree (%) | 51.8 | 49.5 | 41.8 | 52.6 |
| A-level (%) | 40.3 | 40.2 | 40.0 | 46.3 |
| Employed (%) | 66.2 | 59.3 | 62.6 | 63.6 |
| Insured in SHI (%) | 79.5 | 81.0 | 85.3 | 81.0 |
| West Germany (%) | 62.4 | 63.0 | 56.0 | 54.5 |
| BMI (kg/m), mean (SD) | 31.9 (0.3) | 31.5 (0.3) | 28.3 (0.5) | 27.9 (0.5) |
| Glycemic category<sup>a</sup>, mean (SD) | 3.0 (0.1) | 2.9 (0.1) | 2.4 (0.1) | 2.5 (0.1) |
| EQ-5D utility weights (unadjusted), mean (SD) | 0.91 (0.01) | 0.92 (0.01) | 0.90 (0.02) | 0.93 (0.01) |
| Annual costs in € (SHI)<sup>b</sup>, mean (SD) | 2288.97 (485.23) | 2216.49 (172.03) | 1827.27 (256.47) | 1952.62 (272.91) |
| Annual costs in € (society)<sup>c</sup>, mean (SD) | 4403.63 (485.23) | 3720.54 (324.45) | 4640.46 (986.23) | 3144.36 (410.14) |
| *12 months after baseline* |  |  |  |  |
| Number of participants with remission (%) | 111 (31.18) | 86 (24.5) | 52 (52) | 31 (30.69) |
| QALY per participant, mean (SD) | 0.92 (0.00) | 0.92 (0.01) | 0.91 (0.01) | 0.93 (0.01) |
| Annual costs in € (SHI)<sup>2</sup>, mean (SD) | 2692 (423.51) | 2160.74 (324.45) | 2678.20 (425.81) | 1469.97 (231.42) |
| Annual costs in € (society)<sup>2</sup>, mean (SD) | 5507.72 (722.08) | 3917.91 (374.95) | 8180.20 (2097.23) | 3500.22 (697.72) |
| Abbreviations: Intensive lifestyle intervention (HR-INT), conventional lifestyle intervention in high-risk |  |  |  |  |

groups (HR-CONV), conventional lifestyle intervention in low-risk groups (LR-CONV), no lifestyle intervention (LR-CTRL), General Certificate of Education Advanced Level (A-level), body mass index (BMI), EuroQol-5D (EQ-5D), Statutory Health Insurance (SHI), and Standard Deviation (SD)
<sup>a</sup>A value of 2 refers to impaired fasting glucose, and a value of 3 refers to impaired glucose tolerance
<sup>b</sup>Excluding screening costs
<sup>c</sup>Number of participants with remission to normal glucose regulation (i.e., participants without prediabetes or diabetes).

Besides outcomes and costs, we included data such as age, BMI, glycemic category according to the ADA classification (i.e., having impaired fasting glucose, impaired glucose tolerance, or both) (2), sex, marital status, educational status and employment status.

Missing data were handled using multiple imputation (ESM Table 6), assuming data were missing at random, with the number of imputations guided by the percentage of missing observations (25,26). Mean incremental costs and outcomes were estimated using Generalized Linear Models (GLM), adjusting for baseline differences (27). Confidence intervals (CIs) were derived using a non-parametric bootstrap procedure with 1000 iterations (28,29). Cost-effectiveness and cost-utility were assessed using incremental cost-effectiveness ratios (ICER) and incremental cost-utility ratios (ICUR), with results visualized through cost-effectiveness planes and acceptability curves based on a range of willingness-to-pay (WTP) thresholds (30,31). This approach reflects the idea that the payer is generally willing to pay for a remission although there is no consensus on the WTP threshold. In terms of QALY gained, we referred to established WTP thresholds in the international literature (UK: GB£20,000 and GB£30,000 (32); US: a range between US$100,000 - US$150,000 (33)). We used Stata (Version 15.1) to prepare the data and perform subsequent analyses.

### Design of the lifetime-horizon health economic model

Long-term cost-effectiveness was simulated for a cohort of 1000 participants using a four-state Markov model (normoglycemia, prediabetes, diabetes, death) depicted in Figure 1, comparing HR-INT against HR-CONV and LR-CONV against LR-CTRL. Individuals entered the model at the age of 58, corresponding to the average age in the PLIS trial, with existing prediabetes and were followed in 1-year cycles until the age of 90. Individuals could move between states: prediabetes to normoglycemia (i.e. remission) or diabetes or death, normoglycemia to prediabetes or death, and diabetes to death. Transitions from diabetes to prediabetes were not modelled because sustained reversal of established diabetes is rare outside of intensive clinical programs. Death state was the final, absorbing state.

**Fig. 1.**
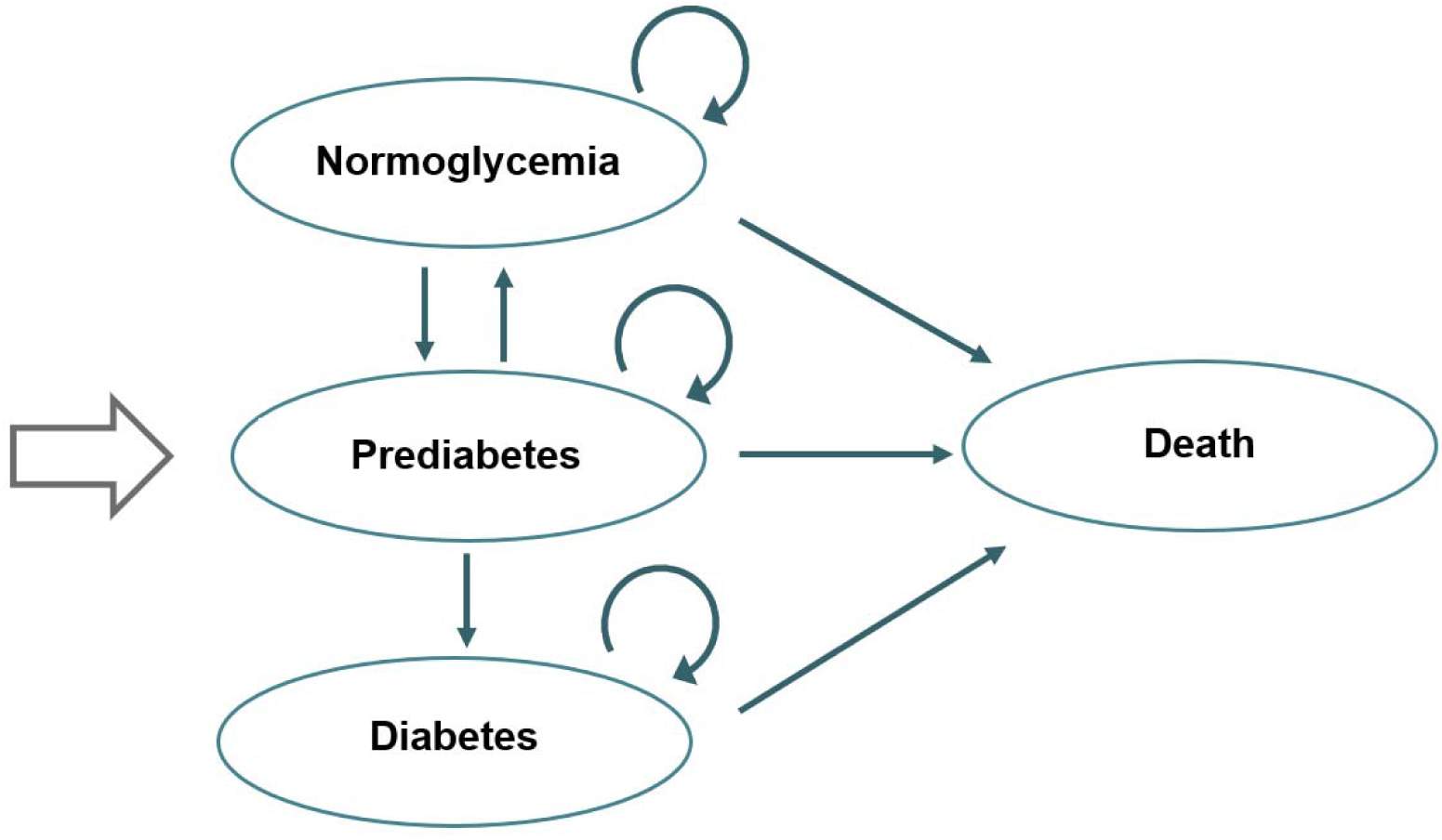
4-state Markov Model

Parameterization of the Markov model involved synthesizing diverse data sources to inform long-term effects. A literature search was conducted to identify suitable data for input parameters, prioritizing comparable modeling studies to guide assumptions and verify data sources. All relevant information was systematically compiled in tables, followed by the selection of appropriate parameter values. Three systematic reviews (34–36) and additional hand searches focusing on economic modeling of prediabetes and diabetes prevention served as a basis.

Model input parameters can be found in Table 1. Transition probabilities between states were taken from the PLIS trial (prediabetes to normoglycemia) and the literature (37–39). For the transitions to the absorbing state “death”, mortality rates were used from German life tables for the general population (40). Hazard ratios were added to model the increased disease-specific probability of dying based on previous diabetes models (39,41).

Costs were calculated from the SHI perspective based on trial data for the prediabetes state (ESM Tables 3 and 4). Drawing upon established modeling approaches, the cost associated with the normoglycemic state was determined to be approximately 38% of the costs attributed to the diabetes state, which was based on nationwide health insurance data from Germany (39,42). Health-related quality of life was measured using average utility values from PLIS for the prediabetes state and the literature (37,43). The analyses were carried out from a healthcare perspective of German SHI as the payer of such a prediabetes program. The cost of lifestyle interventions was calculated based on trial data (ESM Table 5) and applied in the first year. A separate scenario analysis investigated the impact of an adapted costing model where reimbursement was based on outpatient nutritional-therapeutic intervention rates (ESM Table 7), reflecting a structure closer aligned with potential real-world implementation of PLIS interventions.

To be conservative, intervention effects were only applied in year 1, corresponding to the intervention duration. From year 2 onwards, the baseline transition probability for prediabetes remission was used (39). A separate scenario analysis investigated the impact of a sustained intervention effect extending for five years. The discount rate was set at 3% for costs and benefits. Probabilistic sensitivity analysis was performed using 1000 iterations of second-order Monte Carlo simulations. Primary estimates of the base-case were based on probabilistic analysis with 95% CI (2.5th and 97.5th percentiles of distributions) obtained (44). The Markov model was developed in R (Version 4.5.0).

## Results

### Descriptive Analysis

Table 2 shows the characteristics of the population at baseline and 12 months after baseline. Participants were mostly female (about 60%) and had a mean age of approximately 58 years. In the high-risk group, 31% of participants in the intensive intervention achieved remission, compared to 25% in the conventional intervention. In the low-risk group, remission rates were notably higher in the conventional intervention group at 52% compared to 31% in the control group.

The distribution of mean per capita costs at baseline and 12 months after baseline was similar in all study groups (ESM Figure 1).

### Within-trial evaluation

The mean additional costs from the SHI and societal perspective, the number of additional participants with remission (ICER) and the incremental QALYs (ICUR) for the HR-INT and LR-CONV versus the comparators are shown in Table 3a. The ICER was €7081.48 (95% CI: dominated to 47,276.62) and €14,526.92 (95% CI: dominated to 96,891.58) from the perspective of the SHI and society for HR-INT versus HR-CONV, respectively. In the cost-utility analysis, HR-INT was dominated by HR-CONV, indicating that HR-INT incurred higher costs while generating fewer QALYs. For LR-CONV versus LR-CTRL, the ICER was €4278.36 (95% CI: 1312.21 to 11,793.41) and €7007.71 (95% CI: 133.04 to 19,704.88) from the perspective of the SHI and society, respectively. The ICUR was roughly €2.6 million and €4.2 million from the perspective of the SHI and the society, respectively.

**Table 3a.** Results of the within-trial analysis of 100 participants.

| Perspective | Mean incremental outcomes |  | Mean incremental costs<sup>a</sup> | ICER | ICUR |
| --- | --- | --- | --- | --- | --- |
|  | participants with remission<sup>b</sup><br>(95% CI) | QALYs<br>(95% CI) | €<br>(95% CI) | €/participant with remission<br>(95% CI) | €/QALY |
| *HR-INT vs. HR-CONV* |  |  |  |  |  |
| SHI | 6.91<br>(-0.18; 13.60) | -0.08<br>(-0.95; 0.81) | 48,933.00<br>(210.00;<br>93,629.00) | 7081.48<br>(dominated;<br>47,276.62) | dominated |
| Society | 6.91<br>(-0.18; 13.60) | -0.08<br>(-0.95; 0.81) | 100,381.00<br>(9856.00;<br>187,604.00) | 14,526.92<br>(dominated;<br>96,891.58) | dominated |
| *LR-CONV vs. LR-CTRL* |  |  |  |  |  |
| SHI | 23.85<br>(9.10; 40.55) | 0.04<br>(-2.07; 1.85) | 102,039.00<br>(38,777.00;<br>163,514.00) | 4278.36<br>(1312.21;<br>11,793.41) | 2,550,975.00<br>(dominated;<br>1,153,629.00) |
| Society | 23.85<br>(9.10; 40.55) | 0.04<br>(-2.07; 1.85) | 167,134.00<br>(2424.00;<br>300,891.00) | 7007.71<br>(133.04;<br>19,704.88) | 4,178,350.00<br>(dominated;<br>2,052,882.00) |

As shown in Figure 2, most mean incremental cost-outcome pairs were plotted on the north-east quadrant of the cost-effectiveness plane, highlighting the fact that although HR-INT and LR-CONV tend to be more effective in terms of participants with remission, they also tend to be costlier from both perspectives, compared to their comparators. Based on the cost-effectiveness acceptability curve, the probability of HR-INT being cost-effective was approximately 70% at €9809.81; 80% at €13,013.01; and 90% at €22,122.12 per additional participant with remission from the SHI perspective (ESM Figure 2). From the societal perspective, the respective probabilities were achieved at €19,919.92; €26,326.33; and €48,348.35. LR-CONV achieved the same probabilities of cost-effectiveness at €4804.81; €5605.61; and €7407.41 from the SHI perspective and €8308.31; €9809.81; and €12,612.61 from the societal perspective.

**Fig. 2.**
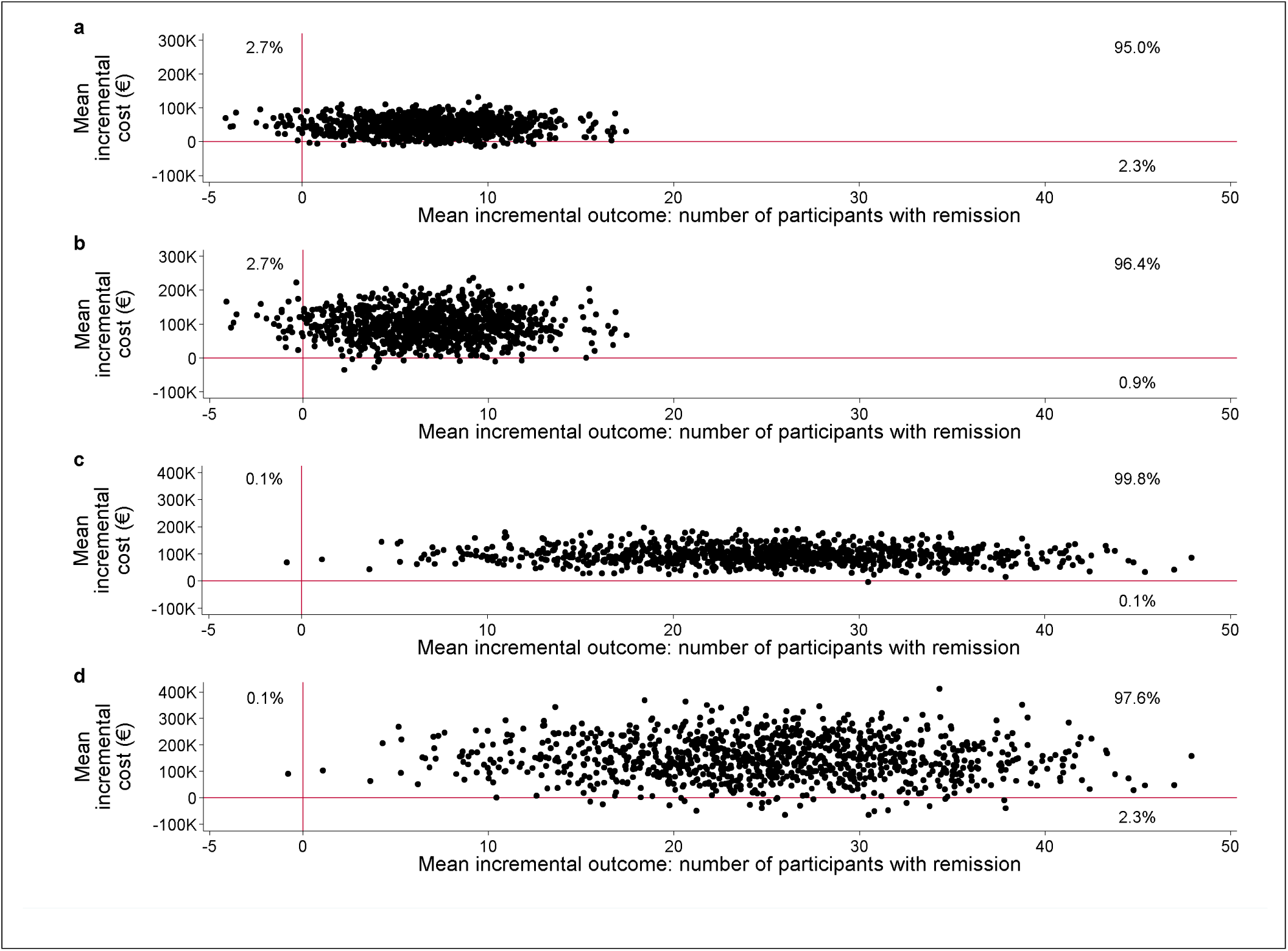
Cost-effectiveness planes of the within-trial evaluation for 100 participants Abbreviations: Incremental cost-effectiveness ratio (ICER), Statutory Health Insurance (SHI), intensive lifestyle intervention (HR-INT), conventional lifestyle intervention in high-risk groups (HR-CONV), conventional lifestyle intervention in low-risk groups (LR-CONV), and no lifestyle intervention (LR-CTRL). Mean incremental costs are presented in thousands of euros. Panel a: HR-INT vs. HR-CONV from the SHI perspective (approx. 95% of incremental cost-outcome pairs in the north-east quadrant). Panel b: HR-INT vs. HR-CONV from the societal perspective (approx. 96% of incremental cost-outcome pairs in the north-east quadrant). Panel c: LR-CONV vs. LR-CTRL from the SHI perspective (almost 100% of incremental cost-outcome pairs in the north-east quadrant). Panel d: LR-CONV vs. LR-CTRL from the societal perspective (approx. 98% of incremental cost-outcome pairs in the north-east quadrant).

In terms of QALYs gained, we did not conclude the probability of HR-INT and LR-CONV being cost-effective because only half or even less of the mean incremental cost-outcome pairs were plotted on the north-east quadrant of the cost-utility plane (HR-INT versus HR-CONV: 41% to 42% from both perspectives; LR-CONV versus LR-CTRL: 51% to 52% from both perspectives) (ESM Figure 3). Additionally, the ICUR of LR-CONV exceeded established WTP thresholds (cost-effectiveness acceptability curves are plotted in ESM Figure 4).

**Fig. 3.**
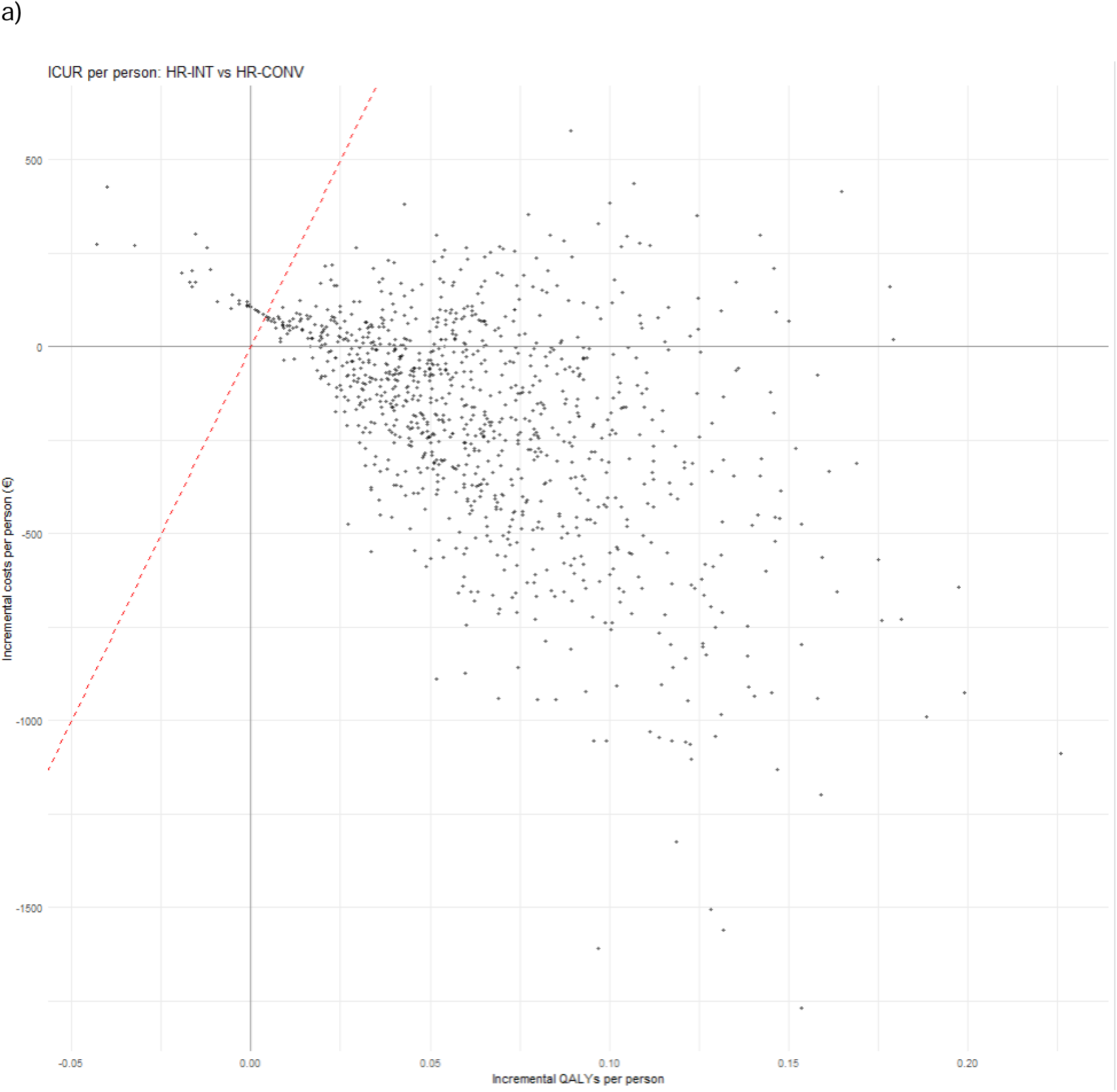
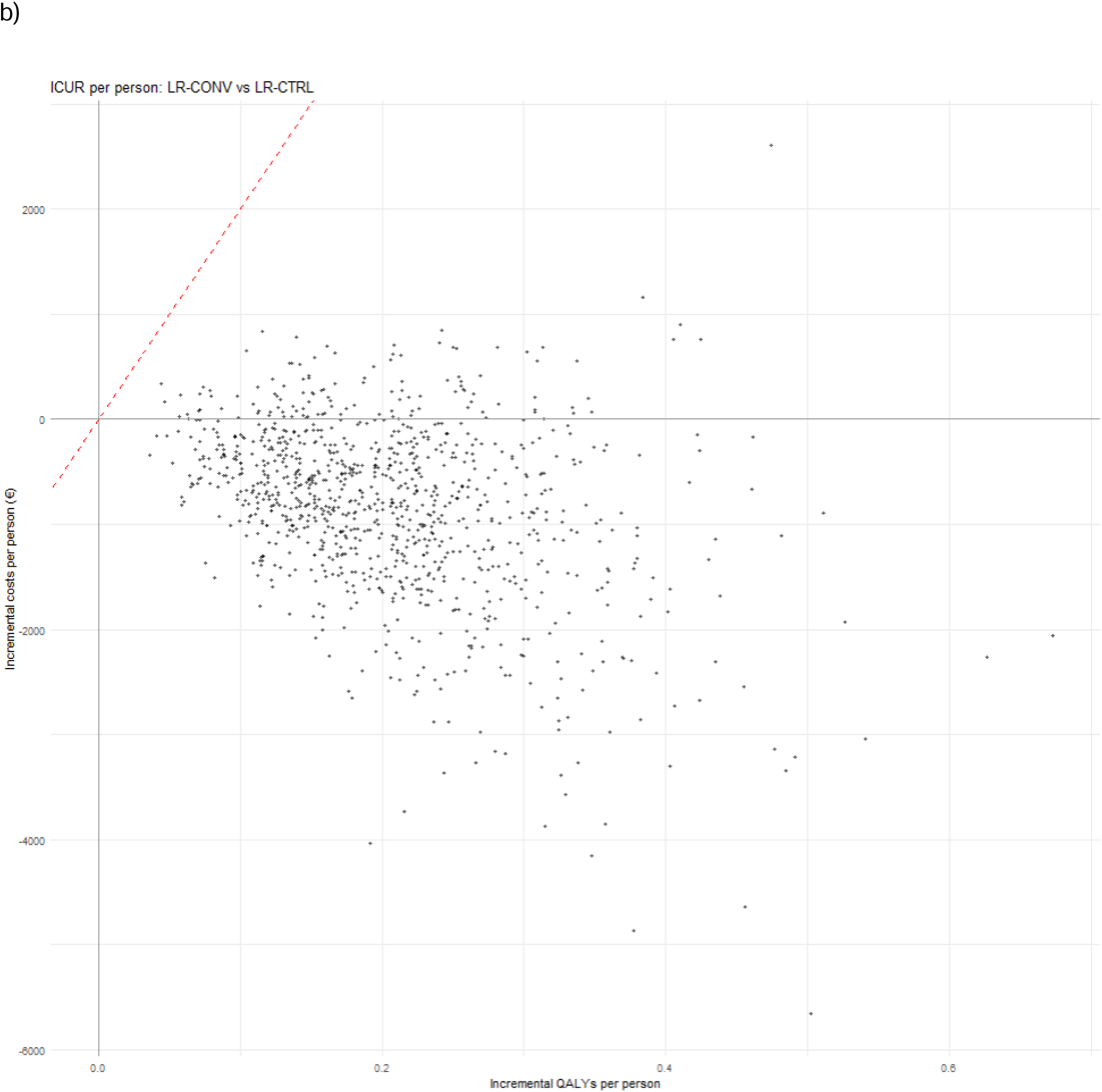
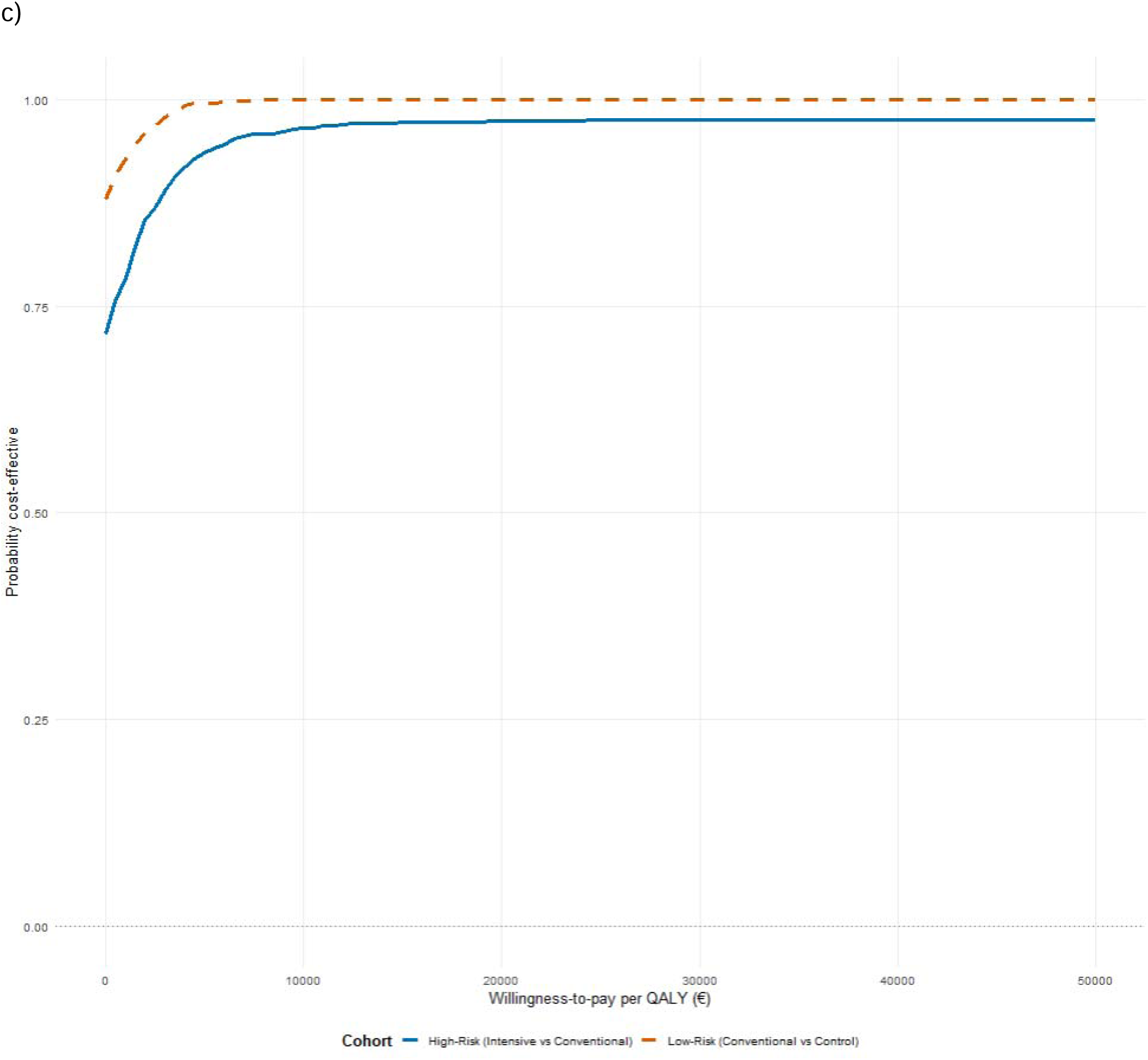
Cost-effectiveness planes and cost-effectiveness acceptability curves of the modeling evaluation Abbreviations: Incremental cost-utility ratio (ICUR), intensive lifestyle intervention (HR-INT), conventional lifestyle intervention in high-risk groups (HR-CONV), conventional lifestyle intervention in low-risk groups (LR-CONV), and no lifestyle intervention (LR-CTRL), quality-adjusted life years (QALYs). Mean incremental costs are presented in euros. Panel a: ICUR per person comparing HR-INT vs. HR-CONV with a hypothetical willingness to pay threshold of 20,000 EUR (red diagonal line) Panel b: ICUR per person comparing LR-CONV vs. LR-CTRL with a hypothetical willingness to pay threshold of 20,000 EUR (red diagonal line) Panel c: Probability of cost-effectiveness for high-risk and low-risk cohorts at different willingness to pay thresholds

### Lifetime simulation model

Base case simulations indicate that both HR-INT and LR-CONV are more effective and less costly over the lifetime time horizon, relative to HR-CONV and LR-CTRL, respectively. While the mean costs decreased by 192€ (95% CI: -926 to 258) and 853€ (95% CI: -2868 to 534) in the HR and LR interventions per participant, respectively, the incremental QALYs gained were estimated at 0.065 (95% CI: 0.004-0.146) and 0.206 (95% CI: 0.071-0.406) per participant (Table 3b). Since the original utility for prediabetes (Table 2) was higher than that for normoglycemia in other studies, the utility for normoglycemia was set one percentage point higher than that for prediabetes, as prediabetes should not plausibly have a higher utility than normoglycemia. The incremental number of participants with prediabetes remission is around 64 for the HR-interventions and 203 for the LR-interventions. Further, the distribution of states is shown in ESM Figure 5.

**Table 3b.** Results of the modeling analyses Base-case

|  | HR-INT | HR-CONV | Increment | LR-CONV | LR-CTRL | Increment |
| --- | --- | --- | --- | --- | --- | --- |
| Cost per participant, € | 53,159 (41,504 - 68,166) | 53,350 (41,466 - 68,706) | -192 (-926 - 258) | 52,097 (41,314 - 65,924) | 52,951 (41,341 - 67,582) | -853 (-2868 - 534) |
| QALY per participant | 12.112 (11.284 - 12.895) | 12.047 (11.167 - 12.841) | 0.065 (0.004 - 0.146) | 12.314 (11.491 - 13.053) | 12.108 (11.254 - 12.876) | 0.206 (0.071 - 0.406) |
| Cost per QALY gained | Cost saving: HR-INT dominates (cost-saving to cost-effective) |  |  | Cost saving: LR-CONV dominates (cost-saving to cost-effective) |  |  |

5-year effect
|  | HR-INT | HR-CONV | Increment | LR-CONV | LR-CTRL | Increment |
| --- | --- | --- | --- | --- | --- | --- |
| Cost per participant, € | 51,663 (41,072 - 65,239) | 52,028 (41,200 - 65,825) | -364 (-1478 - 373) | 50,489 (40,242 - 62,937) | 51,483 (40,696 - 64,754) | -993 (-3346 - 586) |
| QALY per participant | 12.486 (11.672 - 13.249) | 12.378 (11.532 - 13.158) | 0.108 (0.006 - 0.231) | 12.715 (11.935 - 13.474) | 12.476 (11.644 - 13.247) | 0.239 (0.08 - 0.467) |
| Cost per QALY gained | Cost saving: HR-INT dominates (cost-saving to cost-effective) |  |  | Cost saving: LR-CONV dominates (cost-saving to cost-effective) |  |  |

Abbreviations: Statutory Health Insurance (SHI), incremental cost-effectiveness ratio (ICER), incremental cost-utility ratio (ICUR), quality-adjusted life years (QALYs), confidence interval (CI), intensive lifestyle intervention (HR-INT), conventional lifestyle intervention in high-risk groups (HR-CONV), conventional lifestyle intervention in low-risk groups (LR-CONV), and no lifestyle intervention (LR-CTRL).
Regression was performed with control variables; 95% CIs were obtained from the GLM except for the 95% CIs of ICER/ICUR estimates, which resulted from the bootstrap procedure.
<sup>a</sup>Excluding screening costs.
<sup>b</sup>Number of additional participants with remission to normal glucose regulation (i.e., participants without prediabetes or diabetes).

As shown in Figure 3, the cost-effectiveness plane with 1000 Monte Carlo iterations showed that most simulations were plotted in the south-east quadrant (71.7% for HR and 88% for LR), indicating that the HR-INT and LR-CONV were cost-saving, relative to HR-CONV and LR-CTRL, respectively. In the HR cohort, 26.1% of simulations fell in the north-east quadrant, suggesting greater effectiveness at higher cost, and only 2.2% fell in the north-west quadrant.

When a 5-year effect is assumed, the dominance of the intervention strategies becomes more pronounced. HR-INT against HR-CONV shows an increased cost saving of €364 (95% CI: -1478 to - €373) and a higher QALY gain of 0.108 (95% CI: 0.006 to 0.231). For the LR cohort, LR-CONV against LR-CTRL demonstrates a cost saving of €993 (95% CI: -3346 to 586) and a greater QALY gain of 0.239 (95% CI: 0.08 to 0.467). When intervention costs were recalculated using an outpatient costing framework, incremental costs moved closer to cost neutrality while remaining favorable (ESM Table 9, ESM Figure 6).

## Discussion

This health economic evaluation represents the first within-trial and lifetime-horizon modeled cost-effectiveness and cost-utility analysis of lifestyle interventions with different intensities to achieve remission in a population composed of individuals with either a high-risk or low-risk prediabetes phenotype. It is based on the stratification between the LR and HR phenotype, defined by pathophysiological features of T2DM and has previously been described (9,45,46). Impaired insulin secretion and insulin resistance, which define this phenotype, are the key pathophysiological mechanisms driving the development of T2DM.

To discuss the *within-trial cost-effectiveness* concerning participants with remission, we focus on the cost-effectiveness acceptability curves for both interventions, HR-INT (versus HR-CONV) and LR-CONV (versus LR-CTRL) since no willingness-to-pay thresholds for an additional person with remission exists. The cost-effectiveness acceptability curves show that cost-effectiveness with a probability of 90% would be achieved if the payer were willing to pay around €22,100 per additional participant with remission and €48,300 for HR-INT versus the comparator and around €7,400 and €12,600 for LR-CONV versus the comparator, from the perspective of SHI and society, respectively. Furthermore, better cost-effectiveness would be achieved from the SHI than from the societal perspective. Particularly, indirect costs in the form of patient time costs and loss of productivity are drivers of this result.

Our outcome measure of incremental costs per additional person with remission is not directly comparable to the outcome of other studies that analyze lifestyle interventions in people with prediabetes using diabetes cases prevented as an outcome measure. For example, in the US Diabetes Prevention Program, lifestyle intervention versus no lifestyle intervention was associated with costs of US$17,161 from the health-system perspective and US$25,932 from the societal perspective per diabetes case prevented over 3 years (47).

While HR-INT and LR-CONV yield more people with remission to normal glucose regulation, this result does not translate into QALY gains after 12 months. In the within-trial analysis, in most cases, the lifestyle intervention was either dominated by or not likely to be cost-effective when QALYs served as outcome measure considering established international willingness-to-pay thresholds with substantial amount of uncertainty. The cost-effectiveness planes of ICUR are centered around zero, making positive and negative incremental QALYs almost equally likely, with slightly more negative values for HR-INT and slightly more positive values for LR-CONV.

We have two explanations for the lack of change in QALYs: first, it is doubtful whether there is enough room for QALY improvement in a population that is usually free of symptoms and, therefore, considered healthy. Although participants with prediabetes were at risk of developing diabetes (independently of risk phenotype), they were in good health judged by the high baseline value of their utility weight, potentially suggesting a ceiling effect (Table 2). A systematic review indicates that the responsiveness of EQ-5D instruments in detecting health improvements is poorly observed in people with mild health conditions (48). Participants tend to report full health, as observed in conditions such as diabetes (49), and this tendency varies depending on the choice of instrument (50). The second explanation pertains to the short study period of 12 months. Avoiding or delaying the onset of (pre-)diabetes may yield long-term QALY gains, primarily through the prevention of diabetes-related complications, thereby improving overall cost-effectiveness.

For this reason, we developed a *simulation model to evaluate the long-term cost-effectiveness* of the interventions. Our simulation model indicates that both HR-INT and LR-CONV may be cost-saving over the lifetime time horizon from a SHI perspective. Intensification of lifestyle interventions seems to increase rates of prediabetes remission as well as delay and reduce the onset of T2DM. Although QALY gains might be minor on an individual level, their cumulative potential at the population level could still be significant. While our study models the cost-effectiveness of differently intensified lifestyle interventions across sub-phenotypes for prediabetes remission, the DiRECT trial and its associated modeling work were the first to provide health economic evidence on T2DM remission (51,52).

Other modeling studies with a 50-year time horizon (11) or a lifetime horizon (10) analyzing lifestyle interventions with different intensities to prevent diabetes in high-risk individuals without diabetes or with prediabetes (independently of phenotype) found ICURs ranging from €50 (≈GB£44) (11) to €10,187 (≈GB£9,027) (10) (exchange rate: GB£1 = €1.12849 as of 29^th^ December 2017 (53)). These studies concluded that lifestyle interventions were likely to be cost-effective at a WTP threshold of €22,567 (≈GB£20,000; exchange rate GB£1 = €1.12849 as of 29^th^ December 2017 (53)) per QALY. Depending on the subgroup of high-risk individuals (independent of prediabetes and risk phenotype), the lifestyle intervention was even reported to dominate the comparator at a lifetime horizon (10). In our lifetime simulation model, the low-risk subgroup showed greater decreases in costs and higher QALY gains compared to the high-risk subgroup, although this comparison was not the focus of the study. Both subgroups demonstrated cost-saving outcomes, but the low-risk group achieved a larger QALY gain at a lower cost.

Despite increases in remission rates to normoglycemia in the 5-year effect scenario, the resulting QALY gains in both the base case and the scenario analysis remained relatively modest. This is primarily attributed to the similarity in health utility values between the prediabetes and normoglycemia states, suggesting that the perceived health difference between these two conditions is minimal. This limitation is known as the healthy worker effect (54). Consequently, even a prolonged intervention effect leading to more individuals transitioning to normoglycemia yields limited additional QALYs, as prediabetic individuals had already a relatively high quality of life (Table 2).

The strengths of the study include the rigorous analysis of PLIS both from a within-trial and a lifetime-horizon modeled perspective. The health economic evaluation is the first to provide results on the cost-effectiveness and cost-utility for prediabetes remission of two lifestyle interventions with different intensities among participants with different prediabetes risk subphenotypes. Thus, the country’s decision-maker, namely, the Federal Joint Committee (Gemeinsamer Bundesausschuss), which determines the benefit package of the SHI covering about 75 million people, will receive deep insights regarding cost-effectiveness, which depends not only on intervention intensity and the risk phenotype of prediabetes but also on the perspective and time horizon of the analysis. key policy question is whether the PLIS interventions are cost-effective relative to the current standard of care for individuals with prediabetes. Since the health economic evaluation was performed alongside an RCT, outcome and cost data were both collected from the same population under trial-based circumstances. In addition, the (pragmatic) societal perspective provides a detailed overview of additional relevant costs besides intervention costs and costs due to healthcare utilization. The adapted costing scenario which might better reflect reimbursement rates in the outpatient sector showed that the overall cost-effectiveness conclusions are robust.

A limitation of the trial-based health economic evaluation is that outcomes and costs were only observed over a 12-month follow-up, capturing only the short-term impact of lifestyle intervention, as discussed in the literature (7). However, we developed a lifetime Markov model to address this limitation which showcased that QALY gains are seen long-term for the HR-INT and LR-CONV. Although simple Markov cohort state-transition models like ours are widely used and validated, more complex individual-level simulation models could capture physiological variety better but require detailed data that were not available. Furthermore, diabetes-related complications were not explicitly modeled. Instead, average cost and utility values for T2DM were used, likely reflecting a mix of disease severities and some input parameters were derived from non-German settings due to scarcity of relevant data. Since the model assumes stable transition probabilities, this may mask demography-related variation in diabetes remission dynamics over the life course. Importantly, risk stratification in PLIS relied on advanced phenotyping using MRI, which is unlikely to be routinely feasible in standard care, and the costs of such screening procedures were not included in the economic evaluation. Consequently, outcomes should be interpreted conditional on the availability of an efficient and affordable strategy to assess risk among individuals in real-world settings. Indirect costs related to travel and participation time were not assessed in the societal perspective. Lastly, we used a SHI perspective for the modeling exercise as the payer for such a prevention program, omitting the societal perspective from the lifetime analysis, given the retirement age in Germany and the average age of participants in the PLIS trial.

In conclusion, a WTP of around €22,100 and €48,300 for HR-INT versus HR-CONV and €7400 and €12,600 for LR-CONV versus LR-CTRL per additional participant with remission would be required to achieve a 90% probability of cost-effectiveness from the perspective of SHI and society, respectively. After 12 months, intensive lifestyle intervention in people with a high-risk prediabetes phenotype and conventional lifestyle intervention in people with a low-risk prediabetes phenotype seem not to be cost-effective versus their respective comparators in terms of QALYs gained. This absence of short-term QALY gains may reflect both a ceiling effect in a largely asymptomatic population with high baseline utility values and the limited ability of a 12-month observation period to capture downstream benefits of delayed diabetes onset and reduced diabetes-related complications. In the long term, simulation modeling suggests cost-savings from the HR-INT and LR-CONV, relative to HR-CONV and LR-CTRL, respectively, as normoglycemia-associated costs, T2DM-associated costs and utility losses were reduced.

## Acknowledgments

We thank Prof. Dr. Häring, who played a key role in the original development of the PLIS trial. We thank Dr. Burkhard Haastert from Medi Statista in Wuppertal for providing advice on statistical matters. We thank Prof. Dr. Michael Laxy for his important contributions and thoughtful feedback during the conceptualization and interpretation of this work. We also thank the Institute of Health Economics and Health Care Management in Munich for sharing knowledge with us on working with the WiDO database. In addition, we thank the Munich team for the preparatory work relating to the quality-of-life data and cost data.

## Data Availability

All data requests about the PLIS trial should be made directly to the German Center for Diabetes Research (DZD e.V.).

## Author Contributions

The grant application for PLIS was initiated by A.F. and N.S. The health economic evaluation of PLIS was designed by N.C. and A.I. The plan for the within-trial health economic evaluation was written by J.M., revised by M.V. The plan of the matching algorithm for pharmaceuticals was written by M.V. and revised by V.G., and J.M. The matching-algorithm was performed by V.G. The plan for the modeling exercise was written by A.W. and revised by D.M., M.V., N.K. and A.I. Data were provided by R.W. and R.S. The analyses were performed by J.M., A.W., M.V., D.M. and discussed with V.G., N.C., C.-M.D., N.K. and A.I. The paper was drafted by D.M. and J.M., and revised by A.W., M.V., C.-M.D., N.K., R.W., A.F., M.R., N.S., M.F., K.M., F.E.-F., and A.I. The final paper was read and approved by all the authors. D.M., M.V., J.M., A.W. are the guarantors of this work and, as such, have full access to all the data relevant to the health economic evaluation. Therefore, they take responsibility for the integrity of the data and the accuracy of the data analysis.

## Abbreviations

ADA: American Diabetes Association
a-level: General Certificate of Education Advanced Level
BCA: base case analysis
BMI: body mass index
CHEERS: Consolidated Health Economic Evaluation Reported Standards Statement
CI: confidence interval
DZD: German Center for Diabetes Research
EQ-5D: EuroQol-5D
GLM: Generalized Linear Model
GDP: gross domestic product
HR-CONV: conventional lifestyle intervention in high-risk groups
HR-INT: intensive lifestyle intervention
ICER: incremental cost-effectiveness ratio
ICUR: incremental cost-utility ratio
LR-CONV: conventional lifestyle intervention in low-risk groups
LR-CTRL: no lifestyle intervention
MRS: magnetic resonance spectroscopy
OGTT: oral glucose tolerance test
PLIS: Prediabetes Lifestyle Intervention Study
RCT: randomized-controlled trial
SA: sensitivity analysis
SHI: Statutory Health Insurance
QALY: quality-adjusted life years
WTP: willingness-to-pay

## References

1. Genitsaridi I, Salpea P, Salim A, Sajjadi SF, Tomic D, James S, et al. 11th edition of the IDF Diabetes Atlas: global, regional, and national diabetes prevalence estimates for 2024 and projections for 2050. Lancet Diabetes Endocrinol. 2026 Feb;14(2):149–56. doi:10.1016/S2213-8587(25)00299-2 PubMed PMID: 41412135.

2. American Diabetes Association. Diagnosis and classification of diabetes mellitus. Diabetes Care. 2014 Jan;37 Suppl 1:S81–90. doi:10.2337/dc14-S081 PubMed PMID: 24357215.

3. Gerstein HC, Santaguida P, Raina P, Morrison KM, Balion C, Hunt D, et al. Annual incidence and relative risk of diabetes in people with various categories of dysglycemia: A systematic overview and meta-analysis of prospective studies. Diabetes Research and Clinical Practice. 2007 Dec;78(3):305–12. doi:10.1016/j.diabres.2007.05.004

4. Yokota N, Miyakoshi T, Sato Y, Nakasone Y, Yamashita K, Imai T, et al. Predictive models for conversion of prediabetes to diabetes. Journal of Diabetes and its Complications. 2017 Aug;31(8):1266–71. doi:10.1016/j.jdiacomp.2017.01.005

5. Galaviz KI, Weber MB, Suvada K, Gujral UP, Wei J, Merchant R, et al. Interventions for Reversing Prediabetes: A Systematic Review and Meta-Analysis. American Journal of Preventive Medicine. 2022 Apr;62(4):614–25. doi:10.1016/j.amepre.2021.10.020

6. Gebregergish SB, Hashim M, Heeg B, Wilke T, Rauland M, Hostalek U. The cost-effectiveness of metformin in pre-diabetics: a systematic literature review of health economic evaluations. Expert Review of Pharmacoeconomics & Outcomes Research. 2020 Mar 3;20(2):207–19. doi:10.1080/14737167.2020.1688146

7. Glechner A, Keuchel L, Affengruber L, Titscher V, Sommer I, Matyas N, et al. Effects of lifestyle changes on adults with prediabetes: A systematic review and meta-analysis. Primary Care Diabetes. 2018 Oct;12(5):393–408. doi:10.1016/j.pcd.2018.07.003

8. Birkenfeld AL, Mohan V. Prediabetes remission for type 2 diabetes mellitus prevention. Nat Rev Endocrinol. 2024 Aug;20(8):441–2. doi:10.1038/s41574-024-00996-8 PubMed PMID: 38806698.

9. Stefan N, Staiger H, Wagner R, Machann J, Schick F, Häring HU, et al. A high-risk phenotype associates with reduced improvement in glycaemia during a lifestyle intervention in prediabetes. Diabetologia. 2015 Dec;58(12):2877–84. doi:10.1007/s00125-015-3760-z

10. Breeze PR, Thomas C, Squires H, Brennan A, Greaves C, Diggle PJ, et al. The impact of Type 2 diabetes prevention programmes based on risk-identification and lifestyle intervention intensity strategies: a cost-effectiveness analysis. Diabet Med. 2017 May;34(5):632–40. doi:10.1111/dme.13314

11. Roberts S, Craig D, Adler A, McPherson K, Greenhalgh T. Economic evaluation of type 2 diabetes prevention programmes: Markov model of low- and high-intensity lifestyle programmes and metformin in participants with different categories of intermediate hyperglycaemia. BMC Med. 2018 Dec;16(1):16. doi:10.1186/s12916-017-0984-4

12. Duijzer G, Bukman AJ, Meints-Groenveld A, Haveman-Nies A, Jansen SC, Heinrich J, et al. Cost-effectiveness of the SLIMMER diabetes prevention intervention in Dutch primary health care: economic evaluation from a randomised controlled trial. BMC Health Serv Res. 2019 Nov 11;19(1):824. doi:10.1186/s12913-019-4529-8 PubMed PMID: 31711499; PubMed Central PMCID: PMC6849241.

13. Fritsche A, Wagner R, Heni M, Kantartzis K, Machann J, Schick F, et al. Different Effects of Lifestyle Intervention in High- and Low-Risk Prediabetes: Results of the Randomized Controlled Prediabetes Lifestyle Intervention Study (PLIS). Diabetes. 2021 Dec 1;70(12):2785–95. doi:10.2337/db21-0526

14. Sandforth A, von Schwartzenberg RJ, Arreola EV, Hanson RL, Sancar G, Katzenstein S, et al. Mechanisms of weight loss-induced remission in people with prediabetes: a post-hoc analysis of the randomised, controlled, multicentre Prediabetes Lifestyle Intervention Study (PLIS). Lancet Diabetes Endocrinol. 2023 Nov;11(11):798–810. doi:10.1016/S2213-8587(23)00235-8 PubMed PMID: 37769677.

15. Vazquez Arreola E, Gong Q, Hanson RL, Wang J, Sandforth L, He S, et al. Prediabetes remission and cardiovascular morbidity and mortality: post-hoc analyses from the Diabetes Prevention Program Outcome study and the DaQing Diabetes Prevention Outcome study. Lancet Diabetes Endocrinol. 2026 Feb;14(2):137–48. doi:10.1016/S2213-8587(25)00295-5 PubMed PMID: 41397402.

16. Sandforth A, Arreola EV, Hanson RL, Wewer Albrechtsen NJ, Holst JJ, Ahrends R, et al. Prevention of type 2 diabetes through prediabetes remission without weight loss. Nat Med. 2025 Oct;31(10):3330–40. doi:10.1038/s41591-025-03944-9 PubMed PMID: 41023486; PubMed Central PMCID: PMC12532634.

17. Briggs AH, Claxton K, Sculpher M. Decision modelling for health economic evaluation. Repr. [d. korr. Ausg. von 2007]. Oxford: Oxford Univ. Press; 2011. 237 p. (Handbooks in health economic evaluation series).

18. Schulze MB, Hoffmann K, Boeing H, Linseisen J, Rohrmann S, Möhlig M, et al. An Accurate Risk Score Based on Anthropometric, Dietary, and Lifestyle Factors to Predict the Development of Type 2 Diabetes. Diabetes Care. 2007 Mar 1;30(3):510–5. doi:10.2337/dc06-2089

19. Husereau D, Drummond M, Augustovski F, Briggs A, Carswell C, Caulley L, et al. Consolidated Health Economic Evaluation Reporting Standards 2022 (CHEERS 2022) statement: updated reporting guidance for health economic evaluations. BJOG: An International Journal of Obstetrics & Gynaecology. 2022;129(3):336–44. doi:10.1111/1471-0528.17012

20. Sanders GD, Neumann PJ, Basu A, Brock DW, Feeny D, Krahn M, et al. Recommendations for Conduct, Methodological Practices, and Reporting of Cost-effectiveness Analyses: Second Panel on Cost-Effectiveness in Health and Medicine. JAMA. 2016 Sep 13;316(10):1093. doi:10.1001/jama.2016.12195

21. Glick HA, editor. Economic evaluation in clinical trials. 2. ed. Oxford: Oxford Univ. Press; 2015. 252 p. (Handbooks in health economic evaluation series).

22. Greiner W, Claes C, Busschbach JJV, Graf Von Der Schulenburg JM. Validating the EQ-5D with time trade off for the German population. Eur J Health Econ. 2005 Jun;6(2):124–30. doi:10.1007/s10198-004-0264-z

23. Ludwig K, Graf Von Der Schulenburg JM, Greiner W. German Value Set for the EQ-5D-5L. PharmacoEconomics. 2018 Jun;36(6):663–74. doi:10.1007/s40273-018-0615-8

24. Manca A, Hawkins N, Sculpher MJ. Estimating mean QALYs in trial-based cost-effectiveness analysis: the importance of controlling for baseline utility. Health Economics. 2005 May;14(5):487–96. doi:10.1002/hec.944

25. White IR, Royston P, Wood AM. Multiple imputation using chained equations: Issues and guidance for practice. Statistics in Medicine. 2011 Feb 20;30(4):377–99. doi:10.1002/sim.4067

26. Little RJA, Rubin DB. Statistical analysis with missing data. New York: Wiley; 1987. 278 p. (Wiley series in probability and mathematical statistics).

27. Franklin M, Lomas J, Walker S, Young T. An Educational Review About Using Cost Data for the Purpose of Cost-Effectiveness Analysis. PharmacoEconomics. 2019 May;37(5):631–43. doi:10.1007/s40273-019-00771-y

28. Chaudhary MA, Stearns SC. ESTIMATING CONFIDENCE INTERVALS FOR COST-EFFECTIVENESS RATIOS: AN EXAMPLE FROM A RANDOMIZED TRIAL. Statist Med. 1996 Jul 15;15(13):1447–58. doi:10.1002/(SICI)1097-0258(19960715)15:13%3C1447::AID-SIM267%3E3.0.CO;2-V

29. Barber JA, Thompson SG. Analysis of cost data in randomized trials: an application of the non-parametric bootstrap. Statist Med. 2000 Dec 15;19(23):3219–36. doi:10.1002/1097-0258(20001215)19:23%3C3219::AID-SIM623%3E3.0.CO;2-P

30. Black WC. The CE Plane: A Graphic Representation of Cost-Effectiveness. Med Decis Making. 1990 Aug;10(3):212–4. doi:10.1177/0272989X9001000308

31. Fenwick E, Claxton K, Sculpher M. Representing uncertainty: the role of cost-effectiveness acceptability curves. Health Economics. 2001 Dec;10(8):779–87. doi:10.1002/hec.635

32. Devlin N, Parkin D. Does NICE have a cost-effectiveness threshold and what other factors influence its decisions? A binary choice analysis. Health Econ. 2004 May;13(5):437–52. doi:10.1002/hec.864 PubMed PMID: 15127424.

33. Vanness DJ, Lomas J, Ahn H. A Health Opportunity Cost Threshold for Cost-Effectiveness Analysis in the United States. Ann Intern Med. 2021 Jan;174(1):25–32. doi:10.7326/M20-1392

34. Leal J, Morrow LM, Khurshid W, Pagano E, Feenstra T. Decision models of prediabetes populations: A systematic review. Diabetes Obesity Metabolism. 2019 Jul;21(7):1558–69. doi:10.1111/dom.13684

35. Watson SI, Sahota H, Taylor CA, Chen YF, Lilford RJ. Cost-effectiveness of health care service delivery interventions in low and middle income countries: a systematic review. Glob Health Res Policy. 2018;3:17. doi:10.1186/s41256-018-0073-z PubMed Central PMCID: PMC5992822.

36. Roberts S, Barry E, Craig D, Airoldi M, Bevan G, Greenhalgh T. Preventing type 2 diabetes: systematic review of studies of cost-effectiveness of lifestyle programmes and metformin, with and without screening, for pre-diabetes. BMJ Open. 2017 Nov 15;7(11):e017184. doi:10.1136/bmjopen-2017-017184 PubMed Central PMCID: PMC5695352.

37. Neumann A, Schwarz P, Lindholm L. Estimating the cost-effectiveness of lifestyle intervention programmes to prevent diabetes based on an example from Germany: Markov modelling. Cost Eff Resour Alloc. 2011;9(1):17. doi:10.1186/1478-7547-9-17

38. Tabák AG, Herder C, Rathmann W, Brunner EJ, Kivimäki M. Prediabetes: a high-risk state for diabetes development. The Lancet. 2012 Jun;379(9833):2279–90. doi:10.1016/S0140-6736(12)60283-9

39. Palmer AJ, Tucker DMD. Cost and clinical implications of diabetes prevention in an Australian setting: A long-term modeling analysis. Primary Care Diabetes. 2012 Jul;6(2):109–21. doi:10.1016/j.pcd.2011.10.006

40. Federal Statistical Office - Statistisches Bundesamt (Destatis). Sterbetafel (Periodensterbetafel): Deutschland, Jahre, Geschlecht, Vollendetes Alter. [Internet]. 2023. Available from: Available under: https://www.genesis.destatis.de/genesis//online?operation=table&code=12621-0001&bypass=true&levelindex=0&levelid=1699580651272#abreadcrumb

41. Wong CK, Jiao FF, Siu SC, Fung CS, Fong DY, Wong KW, et al. Cost-Effectiveness of a Short Message Service Intervention to Prevent Type 2 Diabetes from Impaired Glucose Tolerance. J Diabetes Res. 2016;2016:1219581. doi:10.1155/2016/1219581 PubMed Central PMCID: PMC4698777.

42. Kähm K, Laxy M, Schneider U, Rogowski WH, Lhachimi SK, Holle R. Health Care Costs Associated With Incident Complications in Patients With Type 2 Diabetes in Germany. Diabetes Care. 2018 May 1;41(5):971–8. doi:10.2337/dc17-1763

43. Laxy M, Becker J, Kähm K, Holle R, Peters A, Thorand B, et al. Utility Decrements Associated With Diabetes and Related Complications: Estimates From a Population-Based Study in Germany. Value Health. 2021 Feb;24(2):274–80. doi:10.1016/j.jval.2020.09.017 PubMed PMID: 33518034.

44. Wilson ECF. Methodological Note: Reporting Deterministic versus Probabilistic Results of Markov, Partitioned Survival and Other Non-Linear Models. Appl Health Econ Health Policy. 2021 Nov;19(6):789–95. doi:10.1007/s40258-021-00664-2 PubMed PMID: 34258732.

45. Stefan N, Fritsche A, Schick F, Häring HU. Phenotypes of prediabetes and stratification of cardiometabolic risk. Lancet Diabetes Endocrinol. 2016 Sep;4(9):789–98. doi:10.1016/S2213-8587(16)00082-6 PubMed PMID: 27185609.

46. Schmid V, Wagner R, Sailer C, Fritsche L, Kantartzis K, Peter A, et al. Non-alcoholic fatty liver disease and impaired proinsulin conversion as newly identified predictors of the long-term non-response to a lifestyle intervention for diabetes prevention: results from the TULIP study. Diabetologia. 2017 Dec;60(12):2341–51. doi:10.1007/s00125-017-4407-z PubMed PMID: 28840257.

47. Diabetes Prevention Program Research Group. Within-Trial Cost-Effectiveness of Lifestyle Intervention or Metformin for the Primary Prevention of Type 2 Diabetes. Diabetes Care. 2003 Sep 1;26(9):2518–23. doi:10.2337/diacare.26.9.2518

48. Payakachat N, Ali MM, Tilford JM. Can The EQ-5D Detect Meaningful Change? A Systematic Review. PharmacoEconomics. 2015 Nov;33(11):1137–54. doi:10.1007/s40273-015-0295-6

49. Janssen MF, Lubetkin EI, Sekhobo JP, Pickard AS. The use of the EQ-5D preference-based health status measure in adults with Type 2 diabetes mellitus: The use of EQ-5D in Type 2 diabetes. Diabetic Medicine. 2011 Apr;28(4):395–413. doi:10.1111/j.1464-5491.2010.03136.x

50. Jankowska A, Młyńczak K, Golicki D. Validity of EQ-5D-5L health-related quality of life questionnaire in self-reported diabetes: evidence from a general population survey. Health Qual Life Outcomes. 2021 Dec;19(1):138. doi:10.1186/s12955-021-01780-2

51. Xin Y, Davies A, Briggs A, McCombie L, Messow CM, Grieve E, et al. Type 2 diabetes remission: 2 year within-trial and lifetime-horizon cost-effectiveness of the Diabetes Remission Clinical Trial (DiRECT)/Counterweight-Plus weight management programme. Diabetologia. 2020 Oct;63(10):2112–22. doi:10.1007/s00125-020-05224-2 PubMed PMID: 32776237; PubMed Central PMCID: PMC7476973.

52. Lean MEJ, Leslie WS, Barnes AC, Brosnahan N, Thom G, McCombie L, et al. Durability of a primary care-led weight-management intervention for remission of type 2 diabetes: 2-year results of the DiRECT open-label, cluster-randomised trial. Lancet Diabetes Endocrinol. 2019 May;7(5):344–55. doi:10.1016/S2213-8587(19)30068-3 PubMed PMID: 30852132.

53. EXCHANGE-RATES.ORG. US-Dollars (USD) in Euro (EUR) umrechnen [Internet]. [cited 2021 Oct 26]. Available from: https://de.exchange-rates.org/HistoricalRates/E/USD/29.12.2017

54. Kuo S, Ye W, Wang D, McEwen LN, Villatoro Santos C, Herman WH. Cost-effectiveness of the National Diabetes Prevention Program: A Real-world, 2-Year Prospective Study. Diabetes Care. 2025 Jul 1;48(7):1180–8. doi:10.2337/dc24-1110

