## Supplement for "One-year within-trial and lifetime-horizon modeled health economic evaluation of the risk-stratified Prediabetes Lifestyle Intervention Study (PLIS) for prediabetes remission in Germany"

**Electronic Supplemental Material (ESM)**

**ESM Table 1:** Impact Inventory Table

| Sector | Type of impact | Included in this reference case analysis from … perspective | | Notes |
| --- | --- | --- | --- | --- |
|  |  | Perspective of the SHI | Perspective of the society |  |
| Formal Health Care Sector | | | | |
| Health | Health Outcomes |  |  |  |
|  | QALYs based on health-related quality of life (EQ-5D) | ✓ | ✓ |  |
|  | Remission to normal glucose | ✓ | ✓ |  |
|  | Medical Costs |  |  |  |
|  | Costs for providing the intervention:   - training costs for lifestyle advisors - time cost of lifestyle advisors - overhead | ✓ | ✓ | Costs are be paid by SHI if the intervention is transferred to standard care |
|  | Costs for health care utilization paid by SHI   - Outpatient care - Hospital stays - Rehabilitation - Pharmaceuticals | ✓ | ✓ |  |
|  | Costs for rehabilitation paid by German pension insurance | - | ✓ |  |
|  | Out of pocket costs and co-payments for health care utilization paid by patients   - Hospital stays - Rehabilitation - Pharmaceuticals | - | ✓ |  |
| Informal Health Care Sector | | | | |
| Health | Patient-time cost for physical activity | - | ✓ |  |
|  | Out of pocket costs (e.g. devices for physical activity and commercial fitness programmes) | - | ✓ |  |
| Non-Health Care Sectors | | | | |
| Productivity | Cost of unpaid lost productivity due to absent days from work | - | ✓ |  |

**ESM Table 2:** Mean EQ-5D score and respective EQ-5D dimensions

|  |  | HR-INT | HR-CONV | LR-CONV | LR-CTRL | p-value^a^  HR-INT vs. HR-CONV | p-value^b^  LR-CONV vs. LR-CTRL |
| --- | --- | --- | --- | --- | --- | --- | --- |
| *T0* | *EQ-5D-Score (EQ-5D-3L and EQ-5D-5L)* | | | | | | |
|  | EQ-5D-Score | 0.91 | 0.92 | 0.90 | 0.93 | 0.012 | 0.444 |
|  | N | 329 | 318 | 88 | 88 |  |  |
|  | *EQ-5D-3L* | | | | | | |
|  | Mobility | 1.18 | 1.07 | 1.14 | 1.05 | 0.133 | 0.334 |
|  | Self-care | 1.03 | 1.00 | 1.00 | 1.00 | 0.294 | / |
|  | Usual Activities | 1.18 | 1.11 | 1.19 | 1.10 | 0.425 | 0.405 |
|  | Pain / Discomfort | 1.63 | 1.43 | 1.67 | 1.43 | 0.078 | 0.079 |
|  | Anxiety / Depression | 1.30 | 1.18 | 1.19 | 1.33 | 0.289 | 0.933 |
|  | N | 40 | 44 | 21 | 21 |  |  |
|  | *EQ-5D-5L* | | | | | | |
|  | Mobility | 1.40 | 1.33 | 1.24 | 1.24 | 0.036 | 0.869 |
|  | Self-care | 1.04 | 1.01 | 1.06 | 1.07 | 0.055 | 0.731 |
|  | Usual Activities | 1.25 | 1.18 | 1.31 | 1.16 | 0.111 | 0.300 |
|  | Pain / Discomfort | 1.92 | 1.84 | 1.82 | 1.73 | 0.088 | 0.897 |
|  | Anxiety / Depression | 1.30 | 1.34 | 1.45 | 1.39 | 0.684 | 0.833 |
|  | N | 289 | 274 | 67 | 67 |  |  |
| *T1* | *EQ-5D-Score (EQ-5D-3L and EQ-5D-5L)* | | | | | | |
|  | EQ-5D-Score | 0.92 | 0.93 | 0.92 | 0.93 | 0.247 | 0.344 |
|  | N | 286 | 261 | 82 | 77 |  |  |
|  | *EQ-5D-3L* | | | | | | |
|  | Mobility | 1.15 | 1.04 | 1.14 | 1.00 | 0.191 | 0.150 |
|  | Self-care | 1.00 | 1.00 | 1.00 | 1.00 |  |  |
|  | Usual Activities | 1.04 | 1.08 | 1.07 | 1.29 | 0.511 | 0.146 |
|  | Pain / Discomfort | 1.48 | 1.36 | 1.50 | 1.57 | 0.380 | 0.710 |
|  | Anxiety / Depression | 1.15 | 1.16 | 1.21 | 1.36 | 0.817 | 0.411 |
|  | N | 27 | 25 | 14 | 14 |  |  |
|  | *EQ-5D-5L* | | | | | | |
|  | Mobility | 1.31 | 1.31 | 1.22 | 1.29 | 0.728 | 0.790 |
|  | Self-care | 1.02 | 1.01 | 1.04 | 1.08 | 0.722 | 0.932 |
|  | Usual Activities | 1.20 | 1.14 | 1.18 | 1.21 | 0.515 | 0.530 |
|  | Pain / Discomfort | 1.83 | 1.77 | 1.85 | 1.52 | 0.295 | 0.019 |
|  | Anxiety / Depression | 1.30 | 1.27 | 1.35 | 1.40 | 0.638 | 0.418 |
|  | N | 259 | 236 | 68 | 63 |  |  |
| *T2* | *EQ-5D-Score (EQ-5D-3L and EQ-5D-5L)* | | | | | | |
|  | EQ-5D-Score | 0.92 | 0.93 | 0.91 | 0.93 | 0.192 | 0.127 |
|  | N | 323 | 316 | 88 | 90 |  |  |
|  | *EQ-5D-3L* | | | | | | |
|  | Mobility | 1.09 | 1.10 | 1.07 | 1.21 | 0.936 | 0.289 |
|  | Self-care | 1.00 | 1.00 | 1.00 | 1.00 | / | / |
|  | Usual Activities | 1.09 | 1.13 | 1.07 | 1.14 | 0.905 | 0.549 |
|  | Pain / Discomfort | 1.61 | 1.58 | 1.71 | 1.36 | 0.762 | 0.063 |
|  | Anxiety / Depression | 1.21 | 1.16 | 1.29 | 1.21 | 0.605 | 0.668 |
|  | N | 33 | 31 | 14 | 14 |  |  |
|  | *EQ-5D-5L* | | | | | | |
|  | Mobility | 1.36 | 1.31 | 1.36 | 1.24 | 0.252 | 0.411 |
|  | Self-care | 1.03 | 1.01 | 1.01 | 1.04 | 0.375 | 0.572 |
|  | Usual Activities | 1.19 | 1.14 | 1.28 | 1.14 | 0.519 | 0.279 |
|  | Pain / Discomfort | 1.78 | 1.71 | 1.88 | 1.66 | 0.263 | 0.076 |
|  | Anxiety / Depression | 1.30 | 1.24 | 1.31 | 1.36 | 0.432 | 0.761 |
|  | N | 290 | 285 | 74 | 76 |  |  |

Abbreviation: Intensive lifestyle intervention (HR-INT), conventional lifestyle intervention in high-risk groups (HR-CONV), conventional lifestyle intervention in low-risk groups (LR-CONV), no lifestyle intervention (LR-CTRL), EuroQol-5D (EQ-5D)

T0: baseline, T1: 6 months after baseline, T2: 12 months after baseline

^a^ p-value of the Mann-Whitney-U-test for differences between HR-INT vs. HR-CONV

^b^ p-value of the Mann-Whitney-U-test for differences between LR-CONV vs. LR-CTRL

**ESM Table 3: Prices categorized by type of costs and perspective in 2017**

| **Cost items** | **Price per unit** | | **Source** |
| --- | --- | --- | --- |
|  | **Perspective of the SHI** | **Perspective of the society** |  |
| Screening |  |  |  |
| MRS | 256·48€/examination | 256·48€/examination | [1] |
| OGTT | 13·58€/test | 13·58€/test | [2] |
| Outpatient medical care |  | | |
| Primary physicians | 64·61€/treatment case | 64·61€/treatment case | [3] |
| Internists | 141·92€/treatment case | 141·92€/treatment case |  |
| Gynecologists | 49·14€/treatment case | 49·14€/treatment case |  |
| Surgeons | 69·30€/treatment case | 69·30€/treatment case |  |
| Orthopedists | 51·59€/treatment case | 51·59€/treatment case |  |
| Neurologists/Psychiatrists | 71·71€/treatment case | 71·71€/treatment case |  |
| Dermatologists | 36·71€/treatment case | 36·71€/treatment case |  |
| Oculists | 55·38€/treatment case | 55·38€/treatment case |  |
| Urologists | 50·14€/treatment case | 50·14€/treatment case |  |
| Psychotherapists | 395·19€/treatment case | 395·19€/treatment case |  |
| Otolaryngologists | 41·61€/treatment case | 41·61€/treatment case |  |
| Dentists | 100·35€/treatment case | 100·35€/treatment case | [4] |
| Hospital | 12·64€/treatment case | 12·64€/treatment case | [2] |
| Inpatient hospital care | 3,714·42€/ treatment case -10·00€/day | 3,714·42€/ treatment case | [5, 6] |
| Rehabilitation |  | | |
| outpatient | 60·28€/day | 61·79€/day | [7-9] |
| inpatient | 150·42€/day | 154·20€/day |  |
| Pharmaceuticals | varying | varying | [10, 6] |
| Self-management | - | varying | Self-stated |
| Intervention time | 22·90€/hour | 22·90€/hour | [11, 12] |
| Participants’ time |  | | |
| men | - | 18·17€/hour | [11-13] |
| women | - | 15·49€/hour |  |
| Productivity loss due to work absence |  | | |
| men | - | 303·77 €/ day | [11, 12, 14] |
| women | - | 248·77 €/day |  |

Abbreviation: Statutory Health Insurance (SHI) Magnet-resonance-spectroscopy (MRS), Oral glucose tolerance test (OGTT)

If accessible, we preferred price data of calendar quarter 3 in year 2017 because the last participant completed the 12-month follow-up in this calendar quarter

**ESM Table 4: Approach to calculate costs**

| **Approach** | **Description** |
| --- | --- |
| **Collecting data** | |
| Healthcare utilization | A self-reported questionnaire based on a validated instrument [15] was used to quantify healthcare utilization associated with outpatient physician visits (including dentist visits), ambulant surgeries, (day) hospital stays, rehabilitation days, and medication intake at baseline and 12 months after baseline with a 6-month recall period (except ambulant physician visits with a 3-month-recall period and medication intake with a 7-day-recall period). We did not consider ambulant surgeries and day hospital stays in the analysis because information was not sufficient.  Information on out-of-pocket payments of participants for self-management, i.e., taking part in commercial programmes or acquiring products to improve health, was collected with the German translation of the self-stated questionnaire of the US Diabetes Prevention Programme (DPP, Resource Utilisation and Cost of Intervention Questionnaire) [16] 6 months and 12 months after baseline with a recall period of 6 months. |
| Intervention-related | Therapy protocols were used to quantify the time of lifestyle advisors spent with participants for phone contacts, check-ups, and for delivering sessions of dietary and exercise counselling. |
| Productivity loss due to work absence (absenteeism) | Absenteeism as days of work absence was quantified using a self-reported questionnaire at baseline and 12 months after baseline with a 6-month recall period. |
| **Quantifying and pricing** | |
| Screening | |
| MRS | Price of MRS referred to magnet resonance tomography (MRT) with service number 5720 of the “Gebührenordung für Ärzte”[1]. According to correspondence with an employee of the private health insurance, price of MRS was similar to MRT. |
| OGTT | Price referred to health service with „Gebührenordnungsposition“ 01777 of the EBM to rule out gestation diabetes [2]. As OGTT to rule out diabetes was not listed in the EBM, we assumed equal prices. |
| Outpatient medical care | Published sources from German statistics do not quantify the number of visits but the number of treatment cases, which is defined as the treatment of the same insured person by the same healthcare provider in a calendar quarter at the expense of the same health insurance company. As the number of visits was quantified in PLIS, we translated visits to treatment cases assuming that one visit equaled one treatment case per calendar quarter, whereas more than one visit equaled two treatment cases. |
| Physician | We did not consider rarely visited physician groups (i.e., if they were visited in less than 1% of total observations).  Price was calculated by dividing the fee turnover by the number of treatment cases [3]. Weighted average was calculated for neurologists/psychiatrists. The fee turnovers of both physicians were weighted with respect to the share of treatment cases. |
| Dentist | Price was calculated by dividing the billed amounts of dental services by the number of dental treatments [4]. |
| Hospital | Price based on an emergency fee (fee item 01210 of the “Einheitlicher Bewertungsmaßstab”(EBM)) [2]. |
| Inpatient hospital care | Price from the SHI perspective based on the revenue volume per treatment case [5] excluding out-of-pocket payments of 10€ per each hospital day [6]. |
| Rehabilitation | We considered only participants who were not employed as these costs would be relevant for the SHI. Price from the SHI perspective was calculated as described in [17] by dividing the outpatient (inpatient) expenditures [8] by the sum of outpatient (inpatient) days used for rehabilitation and prevention [7]. To calculate the price from the societal perspective, first, the share of out-of-pocket payments was determined by dividing the total (outpatient and inpatient) out-of-pocket payments [9] by the total (outpatient and inpatient) expenditures including out-of-pocket-payments. Secondly, out-of-pocket payments were calculated by multiplying the share by the price from the SHI perspective. Thirdly, out-of-pocket payments were summed up with the price from the SHI perspective. |
| Pharmaceuticals | Pharmaceutical prices were calculated based on defined daily dose (DDD) from the December 2017 database of the German Drug Index provided by the AOK Research Institute (WIdO) [10]. We only included pharmaceuticals in the analysis if price information from the database of WIdO could be matched to the dataset of PLIS, either directly via the pharmaceutical name and pharmaceutical registration number (PZN) - a nationwide standardized number to identify pharmaceuticals in Germany, or indirectly via a matching algorithm based on pharmaceutical name, potency, and package size.  Pharmaceutical prices from the SHI perspective were adjusted for out-of-pocket payments and pharmacy discounts but could not be adjusted for other discounts because this information is varying and kept confidential (i.e., pharmacy price minus out-of-pocket payments of participants such as additional costs and copayments, and minus pharmacy discount; discount of the drug manufacturer and discount specific to health insurance companies could not be excluded). Afterwards, the adjusted pharmacy price was divided by the sum of defined daily doses per pharmaceutical package.  Additional costs of participants were defined as the positive differential between pharmacy price and the fixed amount paid by SHIs for a pharmaceutical. Participants either  a) fully pay for pharmaceuticals if the pharmacy price is below €5, or they have copayments of b) €5 if pharmacy price ranges between €5-50 c) 10% of the pharmacy price if it ranges between €51-100, and d) €10 if pharmacy price is higher than €100 [6].  To calculate price from the societal perspective, we summed up adjusted pharmacy price with out-of-pocket payments. |
| Self-management | Prices were self-assessed by participants. |
| Intervention time | The time of lifestyle advisors was evaluated based on gross hourly wages plus social contributions of the employer [18] as we expected these expenses to arise in association with the employment of the workforce.  We assumed lifestyle advisors to be full-time employees working in positions as professionals (corresponding to group 3) in the healthcare sector [11]. We added social security contributions of the employer (9·35% Statutory Pension Insurance; 1·50% Statutory Unemployment Insurance; 7·30% SHI; 1·275% Statutory Care Insurance) to gross hourly wages were [12]. |
| Participants’ time | The time of participants was evaluated based on net hourly wages, as these costs reflected opportunity costs of participants [18].  Participants’ time costs were calculated for men and women assuming them to be full-time employees working in all positions (corresponding to group one to five) in the manufacturing industry and service sector [11]. Net hourly wages of men and women were calculated from gross hourly wages by using the corresponding net share of full-time employed singles [19]. A nation-wide net-share was not available. Therefore, we weighted the net share by the percentage of full-time employees (irrespective of sex) living in the old federal states and in the new federal states [19].  We added contributions paid by employees to the pension insurance 9·35% and to the unemployment insurance 1·5% to net wages of men and women [12] as both serve as potential future income in case of retirement or unemployment [17]. Insurance contributions were calculated from gross hourly wages. |
| Productivity loss due to work absence (absenteeism) | Productivity losses due to absenteeism were calculated according to the human capital approach based on labor costs of a 1 day work absence (i.e., gross hourly wages plus social contributions of the employer) [19].  We assumed men and women to be full-time employees working in all positions (corresponding to group one to five) in the manufacturing industry and service sector [11].  We added social security contributions of the employer (9·35% Statutory Pension Insurance; 1·5% Statutory Unemployment Insurance; 7·3% SHI; 1·275% Statutory Care Insurance) to gross annual earnings of men and women [12].  To calculate absenteeism of 1 day, we divided the adjusted gross annual earnings by the annual working days (excluding sick leave and holidays) [13]. |
| **Calculating costs** | We reported screening costs for the OGTT and the MRS at baseline in the descriptive analysis to inform about the costs of risk-stratification. Despite the importance of screening prior to intervention delivery, we did not include its costs in further analyses since costs should be equal in all groups.  In addition to the costs from the SHI perspective, we added out-of-pocket payments for hospital stays, rehabilitation days, and medication intake in the societal perspective. To keep it pragmatic, we did not consider regulations associated with out-of-pocket payments in the German healthcare system such as ceilings of a maximum payment per year or participants’ eligibility for exemption.  To receive aggregated annual costs, cost components measured retrospectively for shorter periods than a year were linearly extrapolated and summed up. Linear extrapolation was applied neither to pharmaceutical costs as they depended on duration of medication intake nor to self-management costs as the sum of measurements (6 and 12 months after baseline) yielded annual costs. |
| Abbreviation: Statutory Health Insurance (SHI) Magnet-resonance-spectroscopy (MRS), Oral glucose tolerance test (OGTT) | |

ESM Table 5: Intervention related costs

|  |  | HR-INT  (N = 356) | HR-CONV  (N = 351) | LR-CONV  (N = 100) | LR-CTRL  (N = 101) |
| --- | --- | --- | --- | --- | --- |
| *Intervention related costs relevant for SHI perspective and perspective of the society* | | | | | |
| Training cost for live style advisors | | | | | |
|  | Proportional time of live style advisors receiving training^a^ | 1,286.02 € | 773.19 € | 178.79 € | 52.00 € |
|  | Proportional time for providing training^b^ | 221.09 € | 132.93 € | 30.74 € | 8.94 € |
|  | Total | 1,507.11 € | 906.12 € | 209.53 € | 60.94 € |
| Materials | | | | | |
|  | Material required for providing the intervention^c^ | 0.00 € | 0.00 € | 0.00 € | 0.00 € |
| Time of lifestyle advisors to interact with participants and to deliver sessions | | | | | |
|  | Interaction via phone | 194.65 € | 145.03 € | 34.35 € | 55.34 € |
|  | Dietary and activity consultation (h) | 72,081.56 € | 43,378.33 € | 10,159.97 € | 3,005.62 € |
|  | Love and attention | 13,432.76 € | 8,007.37 € | 1,721.32 € | 404.57 € |
|  | Total | 85,708.97 € | 51,530.73 € | 11,915.63 € | 3,465.53 € |
| Overhead | | | | | |
|  | Overhead^d^ | 12,856.35 € | 7,729.61 € | 1,787.34 € | 519.83 € |
| Total intervention related cost (SHI Perspective) | | 100,072.43 € | 60,166.45 € | 13,912.50 € | 4,046.30 € |
| Total intervention related cost per person (SHI Perspective) | | 281.10 € | 171.41 € | 139.13 € | 40.06 € |
| *Intervention related costs relevant for the perspective of the society* | | | | | |
| Patient time cost for physical activity | | 61,566.74 € | 37,617.37 € | 8505.10 € | 2468.79 € |
| Out of pocket costs (e.g. devices for physical activity and fitness) | | 103,620.99 € | 103,820.57 € | 33,130.72 € | 18,549.52 € |
| Total intervention related cost (Perspective of the society)^e^ | | 265,260.17 € | 201,604.39 € | 55,548.32 € | 25,064.61 € |
| Total intervention related cost per person (Perspective of the society) | | 745.11 € | 574.37 € | 555.48 € | 248.16 € |

^a^ In total 10 lifestyle advisors received training (two workshops a 5h). The workshops did not differentiate for the type of lifestyle intervention. Thus, training costs were distributed proportionally per intervention group with respect to the total time of lifestyle advisors to interact with participants and to deliver sessions.

^b^ Calculation is based on university staff with hourly wages of 39.37€. Costs were distributed proportionally per intervention group with respect to the total time of lifestyle advisors to interact with participants and to deliver sessions.

^c^ There are no relevant costs due to use of materials.

^d^ Overhead costs are not recorded within the study. To derive overhead costs, we assumed overhead costs are 15% of costs due to time of lifestyle advisors to interact with participants and to deliver sessions per intervention group.

^e^ Total intervention related cost (SHI Perspective) are included in Total intervention related cost (Perspective of the society)

**ESM Table 6: Imputation model**

Missing data occurred in additional variables (at baseline), utility weights, and costs (at baseline, 6 months after baseline, and 12 months after baseline).

We followed the guidance by Faria et al. [19] to handle missing data in health economic evaluations conducted within RCTs. We applied multiple imputation by chained equations (MICE) [20, 21], as it allowed to deal with different missing data patterns [22].

To keep the number of variables in the imputation model low as suggested by Hardt et al. [23], we included in the imputation model only variables that were also used in the analysis, i.e., costs (at baseline, 6 months after baseline, and 12 months after baseline, except for intervention costs, screening costs, and participants’ time costs), utility weights (at baseline, 6 months after baseline, and 12 months after baseline), glycaemic category (12 months after baseline), and additional variables (including glycaemic category at baseline).

We imputed costs rather than quantities to be able to aggregate cost components and thus, reduce the number of variables entering the imputation model. This required the dataset to have similar missing data patterns across cost components (i.e., having either complete information or no information on costs for each participant) and to have similar recall periods.

Thus, we summed up costs of all ambulant physician groups with each other, costs of hospital treatment with the costs of rehabilitation, and out-of-pocket payments for hospital treatment with out-of-pocket payments for rehabilitation. Costs due to absenteeism and pharmaceuticals, and out-of-pocket payments for pharmaceuticals and self-management were not summed up due to different missing data patterns.

If cost variables had a high number of zero observations, we applied a two-step imputation [24] by imputing first, the probability of cost variables having positive values and second, the cost variable itself conditioned on positive values. All cost variables entered the two-step imputation with one condition except for pharmaceuticals costs (i.e., probability of pharmaceutical costs and probability of out-of-pocket payments having positive values). Out-of-pocket payments for hospital treatment plus rehabilitation was conditioned on positive hospital plus rehabilitation costs. We imputed log-transformed costs in all imputation models as imputed values were more likely to be stable regardless of the amount of missing data [24].

We applied Predictive Mean Matching (pmm) procedures that randomly selected one of three of nearest observed values to replace missing values in costs, utility weights, and additional continuous variables; and a logit model to replace missing values in additional binary variables and in the probability of cost variables.

The same seed of random number generator was specified for each imputation (i.e., 123). The imputation is performed separately according to treatment assignment to take into account possible differences [25]. Variables with missing values were replaced in the order starting from the lowest to the highest percentage of missing values. After each imputation, the imputed variable was eligible to predict another variable with missing values. The imputation model included different sets of prediction variables depending on the variable being imputed.

In the following section, we described more in detail the selection of prediction variables because not all predictors were appropriate and thus, were excluded from the prediction equation:

| **Variable with missing value** | **Which variables to include into the prediction equation?** | **Which variables to exclude from the prediction equation?** |
| --- | --- | --- |
| Measured at baseline | Glycaemic category and all additional variables both without missing values at baseline, and all imputed additional variables.  In case of a two-step imputation, the probability of a cost variable was used as a condition to replace missing values of the (respective) cost variable measured at the same time period. | Variables with missing values or those measured 12 months after baseline when replacing missing values for variables measured at baseline.  Imputed outcome and cost variables were not used to replace missing values of other variables measured at the same time period. |
| Measured at baseline/12 months after baseline | Glycaemic category and all additional variables both without missing values at baseline, and all imputed additional variables.  In case of a two-step imputation, the probability of a cost variable was used as a condition to replace missing values of the (respective) cost variable measured at the same time period.  Imputed outcome and cost variables of previous time periods were only used to replace the missing values of the respective variable measured in later time periods. | Variables with missing values.  Imputed outcome and cost variables were not used to replace missing values of other variables measured at the same time period.  . |

Whenever the selected predictors in the prediction equation did not lead to successful imputation, we approached as explained in the following:

| **Step** | **Approach** |
| --- | --- |
| I | We kept the selected variables in the prediction equation if they were listed among the predictors that were highly correlated with the variable with missing values. This list was predefined prior to the beginning of the imputation and included a number of 20 highly correlated predictors at maximum.  However, if the selected and highly correlated variables have been listed among the variables that have lead to the error of the imputation procedure, we excluded them from the prediction equation.  In case of a two-step imputation, we kept the variables in the prediction equation if they were highly correlated, have not lead to the error, and fulfilled these criteria in both imputation steps. Thus, the set of selected variables in the two-step imputation models was always equivalent. |
| II | If the error still existed, one after another, we excluded variables from the prediction equation starting with the least correlated variable (i.e., variables listed last among the 20 highly correlated predictors).  This step was repeated until the imputation was successful. |

Outcomes and aggregated annual costs were calculated based on imputed datasets.

**ESM Table 7: Prices in 2017 [21] categorized by type of costs from the societal perspective**

| **Cost items** | **Price per unit** |
| --- | --- |
| Outpatient medical care |  |
| Primary physicians | 22.13€/visit |
| Internists | 65.29€/visit |
| Gynecologists | 34.76€/visit |
| Surgeons | 44.68€/visit |
| Orthopedists | 27.00€/visit |
| Neurologists/Psychiatrists | 50.80€/visit |
| Dermatologists | 21.23€/visit |
| Oculists | 44.90€/visit |
| Urologists | 25.90€/visit |
| Psychotherapists | 81.87€/visit |
| Otolaryngologists | 30.12€/visit |
| Dentists | 52.05€/visit |
| Hospital | Not available |
| Inpatient hospital care |  |
| Normal station | 730.57€/day |
| Intensive care unit | 1,581.66€/day |
| Outpatient rehabilitation | 61.56€/day |
| Inpatient rehabilitation | 145.63€/day |

**ESM Table 8: Details of an adapted costing scenario**

An adapted (ambulatory) costing scenario was calculated based on the Contract on the Details of Care with Nutritional-Therapeutic Remedies [27].

The pricing basis is item no. X5003 “Nutritional-Therapeutic Intervention – Individual Treatment (standard service time 60 minutes, of which at least 30 minutes with the person being treated)”.

This service includes carrying out the intervention with the patient as well as preparation and follow-up (including documentation). Both components of the service are combined into a **“standard service time.”**

For PLIS it is assumed that the specified time for delivering the intervention already includes preparation and follow-up and therefore corresponds to the standard service time.

Below the reimbursement rates and co-payments of the currently available contracts for the provision of nutritional-therapeutic remedies (as of 25 September 2025) are shown.

| X5003 | 2021/2022 | 2023 | 2025 | 2026 |
| --- | --- | --- | --- | --- |
| Reimbursement rate (nutritionist) | 67,82 | 73,89 | 79,66 | 81,65 |

The reimbursement rate for 2017 is not retrievable. Therefore, starting from the 2021 value, it is discounted backward to 2017 using a rate of 3%.

$$\text{Reimbursement Nutritionist}_{2017}=67.82/(1+0.03)^{4}=60.26$$

The statutory health insurance can cover the costs partially or fully if the nutritional therapy is provided according to § 43 SGB V and the required certification is available. Patients who require nutritional therapy may receive a written confirmation of medical necessity from their physician. The form also lists diabetes mellitus as an indication. Prediabetes, however, may fall under nutritional counseling, which may not be automatically reimbursed by health insurance.

For our pragmatic approach, we assumed that X5003 would be applicable for reimbursement of nutritional therapy for prediabetes following the PLIS intervention.

Below a full service description for nutritional-therapeutic interventions according to the Contract under § 125 (1) SGB V for Nutritional Therapy is presented, for reference.

Nutritional-Therapeutic Intervention

Definition:

A nutritional-therapeutic intervention includes, in accordance with the *Guideline Terminology in Clinical Nutrition (2013)* of the German Society for Nutritional Medicine and the *German Nutrition Care Process Manual (G-NCP, 2015)*, the following mandatory components:

1. A nutritional-therapeutic assessment
2. A written therapy plan with defined therapy goals
3. Modification of the therapy plan when necessary
4. Selection of the appropriate intervention format
5. Determination of frequency and duration of contacts or interventions
6. Documentation
7. Monitoring and evaluation of outcomes

The intervention must be best suited both for solving the nutritional problem and for the specific user (insured person). The nutritional-therapeutic intervention may take the form of information, explanation, guidance, counseling, and education.

Indication / Impairment:

Rare congenital metabolic diseases and cystic fibrosis where nutritional therapy, as a medical measure (possibly combined with other measures), is strictly required because otherwise severe intellectual or physical impairments or death may occur.

Therapy Goal:

Achieving, stabilizing, and/or maintaining age-appropriate normal metabolic or nutritional parameters, thereby ensuring:

- age-appropriate physical and mental development
- achievement of a stable nutritional status
- prevention or reduction of disease-related consequences
- avoidance of complications
- maintenance of therapy success
- management of comorbidities
- improved life expectancy and participation

The intervention related costs using an adapted (ambulatory) costing scenario are presented below:

|  |  | HR-INT  (N = 356) | HR-CONV  (N = 351) | LR-CONV  (N = 100) | LR-CTRL  (N = 101) |
| --- | --- | --- | --- | --- | --- |
| *Intervention related costs relevant for SHI perspective and perspective of the society* | | | | | |
| Training cost for live style advisors | | | | | |
|  | Proportional time of live style advisors receiving training^a^ | 3,384.09 € | 2,034.61 € | 470.47 € | 136.83 € |
|  | Proportional time for providing training^b^ | 221.09 € | 132.93 € | 30.74 € | 8.94 € |
|  | Total | 3,605.18 € | 2,167.54 € | 501.21 € | 145.77 € |
| Materials | | | | | |
|  | Material required for providing the intervention^c^ | 0.00 € | 0.00 € | 0.00 € | 0.00 € |
| Time of lifestyle advisors to interact with participants and to deliver sessions | | | | | |
|  | Interaction via phone | 512.21 € | 381.65 € | 90.39 € | 145.63 € |
|  | Dietary and activity consultation (h) | 189,678.39 € | 114,147.51 € | 26,735.35 € | 7,909.12 € |
|  | Love and attention | 35,347.51 € | 21,070.91 € | 4,529.54 € | 1,064.59 € |
|  | Total | 225,538.11 € | 135,600.07 € | 31,355.29 € | 9,119.35 € |
| Overhead | | | | | |
|  | Overhead^d^ | 33,830.72 € | 20,340.01 € | 4,703.29 € | 1,367.90 € |
| Total intervention related cost (SHI Perspective) | | 262,974.00 € | 158,107.62 € | 36,559.79 € | 10,633.02 € |
| Total intervention related cost per person (SHI Perspective) | | 738.69 € | 450.45 € | 365.60 € | 105.28 € |

**ESM Table 9: Results of the modeling analysis using an ambulatory costing scenario**

|  | HR-INT | HR-CONV | Increment | LR-CONV | LR-CTRL | Increment |
| --- | --- | --- | --- | --- | --- | --- |
| Cost per participant, € | 53,603  [41,949 - 68,610] | 53,621 [41,736 - 68,977] | -19 [-753 - 431] | 52,317 [41,534 - 66,143] | 53,014 [41,405 - 67,645] | -697 [-2,712 - 690] |
| QALY per participant | 12.112 [11.284 - 12.895] | 12.047 [11.167 - 12.841] | 0.065 [0.004 - 0.146] | 12.314 [11.491 - 13.053] | 12.108 [11.254 - 12.876] | 0.206 [0.071 - 0.406] |
| Cost per QALY gained | Cost saving: HR-INT dominates [cost-saving to cost-effective] | | | Cost saving: LR-CONV dominates [cost-saving to cost-effective] | | |

**ESM Fig. 1: Distribution of costs at baseline (above) and 12 months after baseline (below)**

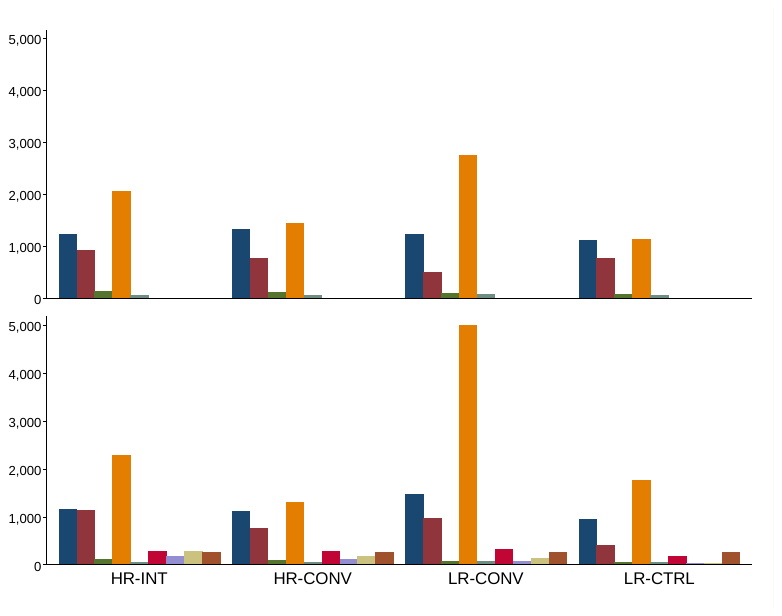

Navy bars= outpatient physician costs, maroon bars= hospital and rehabilitation costs, forest green bars= pharmaceutical costs, dark orange bars= indirect costs, teal bars= out-of-pocket payments, cranberry bars= self-management costs, lavender bars= participants’ time costs, khaki bars= intervention costs, sienna bars= screening costs.

Abbreviation: Statutory Health Insurance (SHI), intensive lifestyle intervention (HR-INT), conventional lifestyle intervention in high-risk groups (HR-CONV), conventional lifestyle intervention in low-risk groups (LR-CONV), no lifestyle intervention (LR-CTRL).

While outpatient physician costs were the largest share of aggregated annual costs from the SHI perspective, indirect costs were highest from the societal perspective. The latter did not seem to be related with the intervention as we observed high indirect costs already at t0. This observation could be coincidental as pointed out in a recent publication [26]Klicken oder tippen Sie hier, um Text einzugeben..

**ESM Fig. 2: Cost-effectiveness acceptability curves for the ICER**

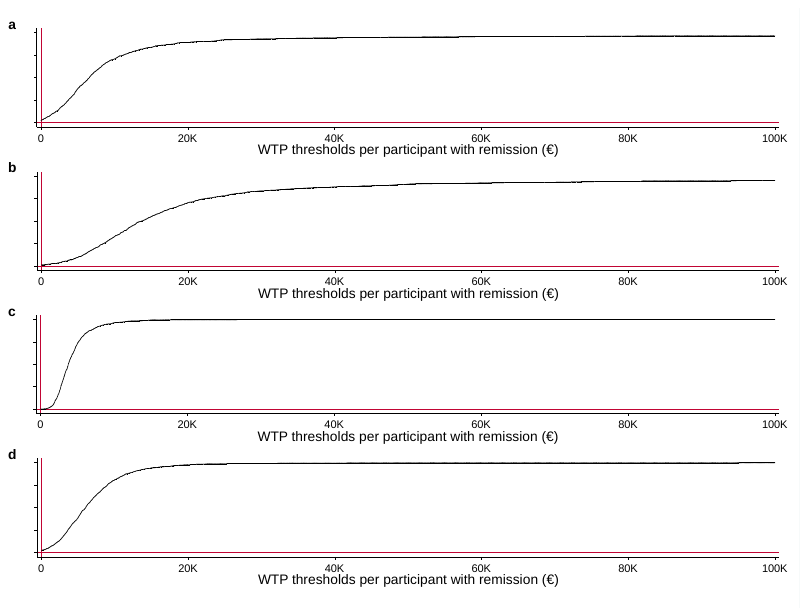

Additional costs per additional participant with remission in euros.

Panel a: HR-INT vs. HR-CONV from the SHI perspective. Panel b: HR-INT vs. HR-CONV from the societal perspective. Panel c: LR-CONV vs. LR-CTRL from the SHI perspective. Panel d: LR-CONV vs. LR-CTRL from the societal perspective.

Abbreviation: Incremental cost-effectiveness ratio (ICER), Statutory Health Insurance (SHI), intensive lifestyle intervention (HR-INT), conventional lifestyle intervention in high-risk groups (HR-CONV), conventional lifestyle intervention in low-risk groups (LR-CONV), no lifestyle intervention (LR-CTRL), willingness-to-pay (WTP).

**ESM Fig. 3: Cost-utility planes of the base case analysis on 100 participants**

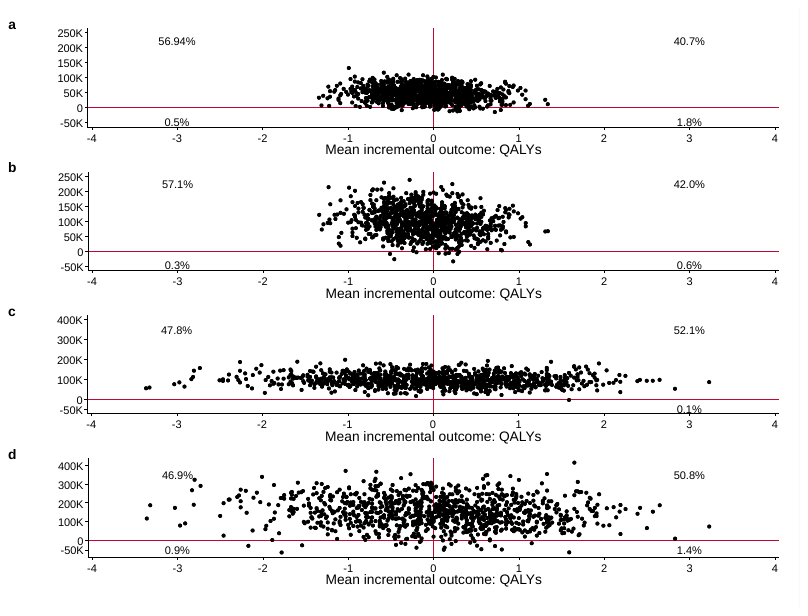

Panel a: HR-INT vs. HR-CONV from the SHI perspective. Panel b: HR-INT vs. HR-CONV from the societal perspective. Panel c: LR-CONV vs. LR-CTRL from the SHI perspective. Panel d: LR-CONV vs. LR-CTRL from the societal perspective.

Abbreviation: Statutory Health Insurance (SHI), quality-adjusted life years (QALY), intensive lifestyle intervention (HR-INT), conventional lifestyle intervention in high-risk groups (HR-CONV), conventional lifestyle intervention in low-risk groups (LR-CONV), no lifestyle intervention (LR-CTRL).

**ESM Fig. 4: Cost-effectiveness acceptability curves for the ICUR**

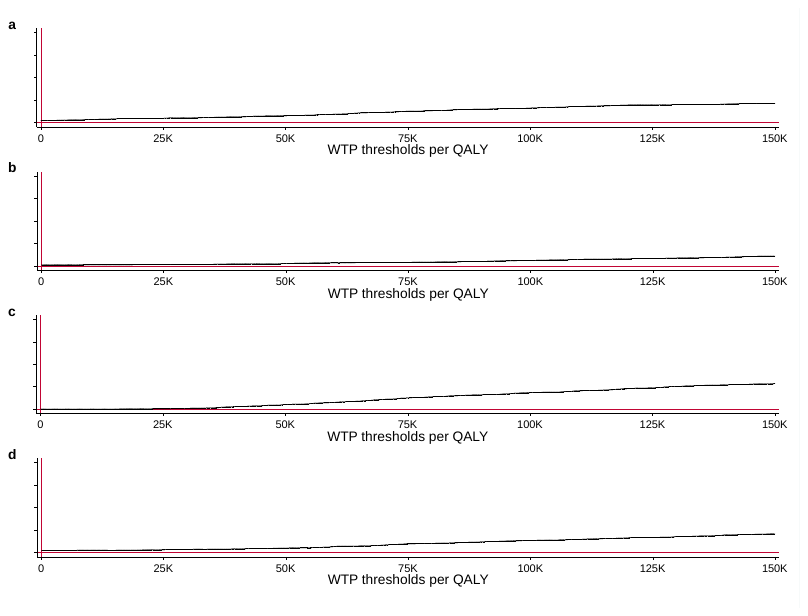

Panel a: HR-INT vs. HR-CONV from the SHI perspective. Panel b: HR-INT vs. HR-CONV from the societal perspective. Panel c: LR-CONV vs. LR-CTRL from the SHI perspective. Panel d: LR-CONV vs. LR-CTRL from the societal perspective.

Abbreviation: Incremental cost-utility ratio (ICUR), Statutory Health Insurance (SHI), intensive lifestyle intervention (HR-INT), conventional lifestyle intervention in high-risk groups (HR-CONV), conventional lifestyle intervention in low-risk groups (LR-CONV), no lifestyle intervention (LR-CTRL), willingness-to-pay (WTP).

**ESM Fig. 5: Distribution of states**

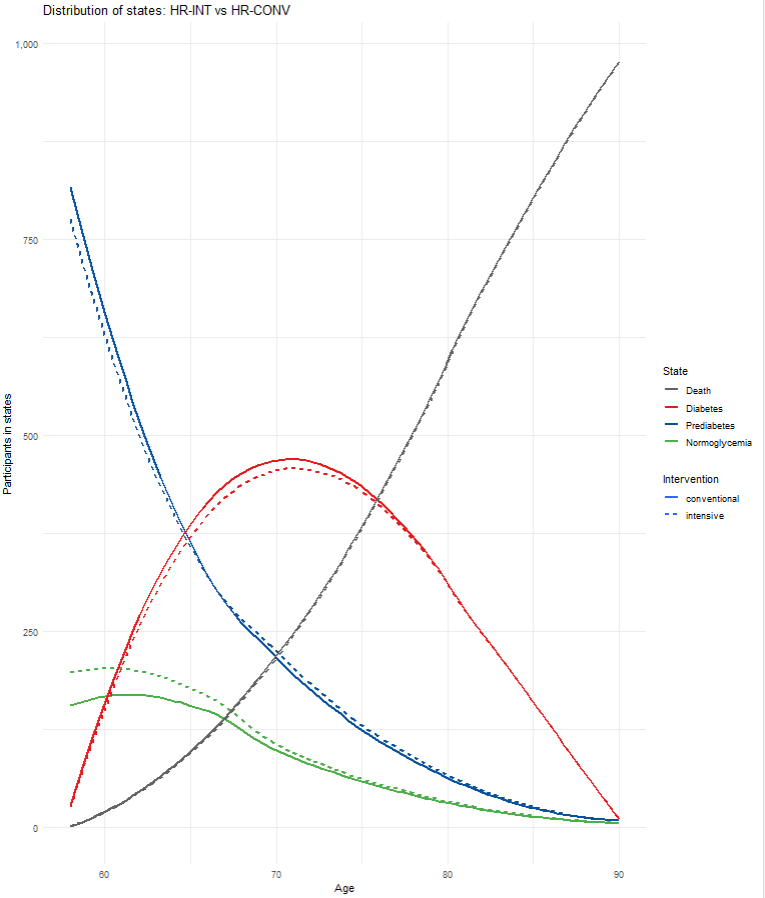

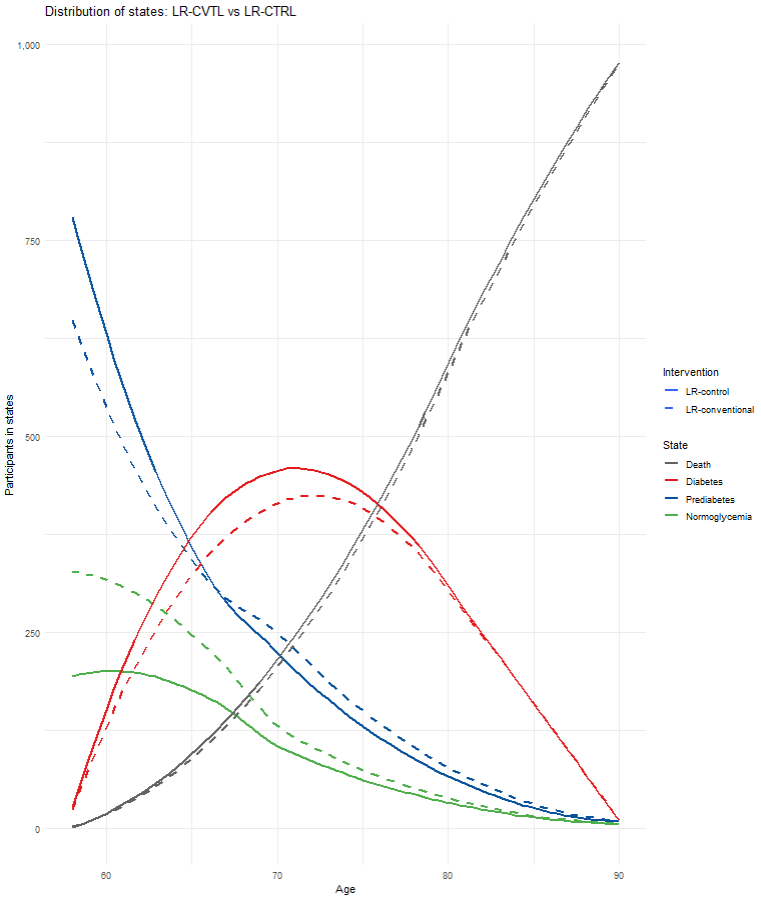

**ESM Fig. 6** Cost-effectiveness planes and cost-effectiveness acceptability curves of the modeling evaluation using an adapted ambulatory costing scenario

a)

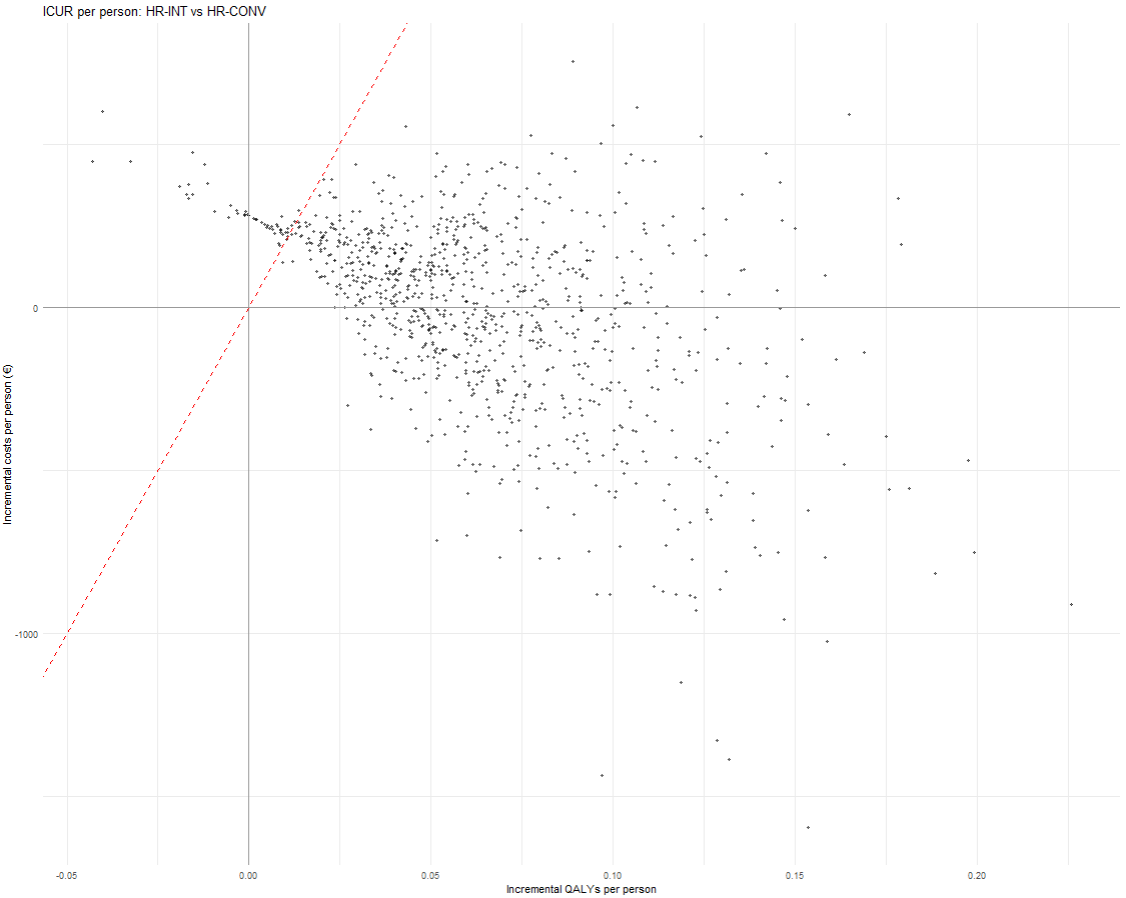

b)

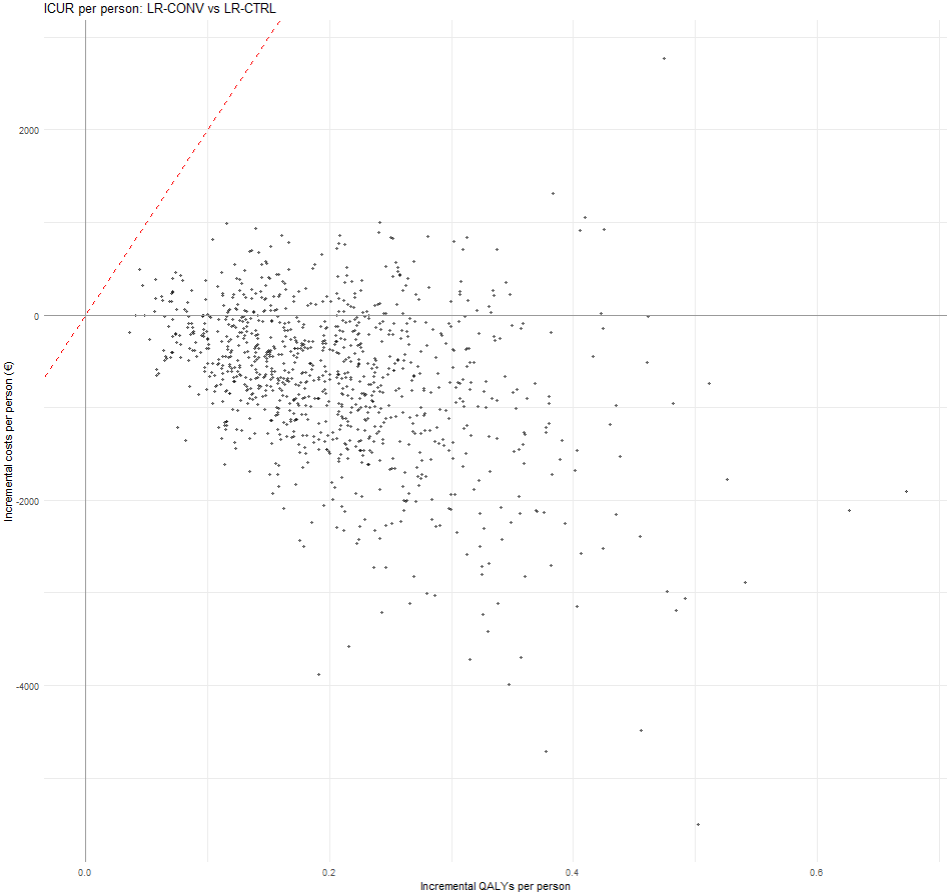

c)
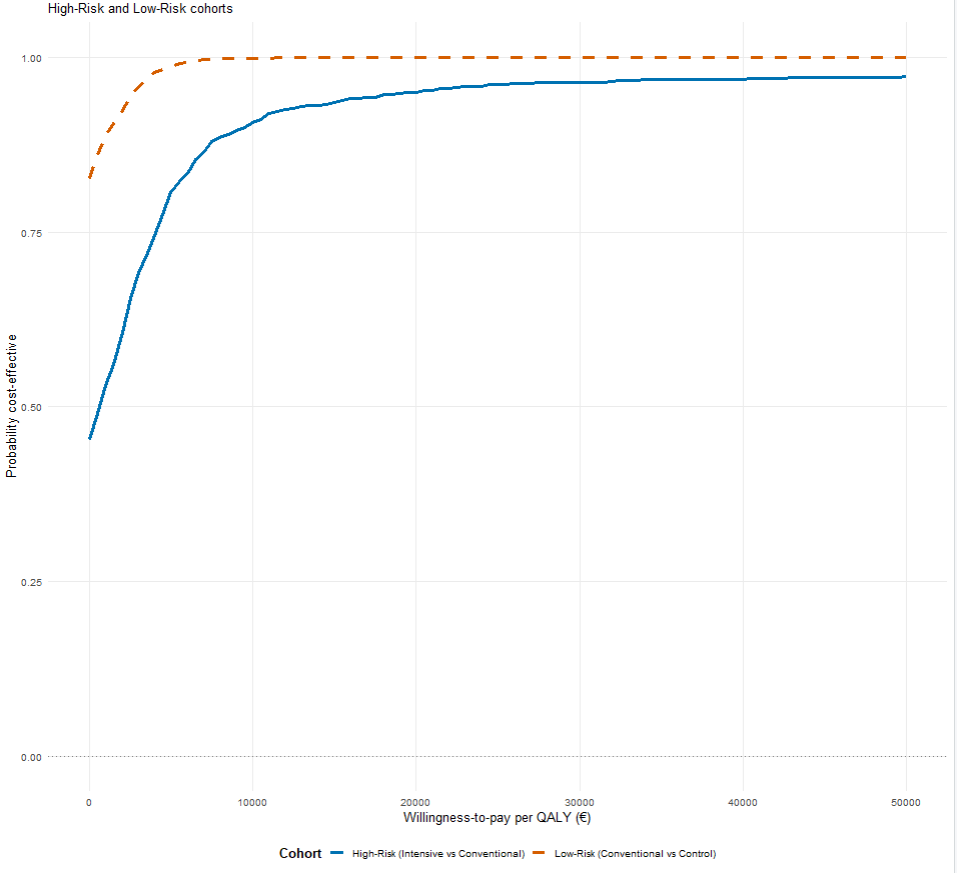

Abbreviations: Incremental cost-utility ratio (ICUR), intensive lifestyle intervention (HR-INT), conventional lifestyle intervention in high-risk groups (HR-CONV), conventional lifestyle intervention in low-risk groups (LR-CONV), and no lifestyle intervention (LR-CTRL), quality-adjusted life years (QALYs).

Mean incremental costs are presented in euros.

Panel a: ICUR per person comparing HR-INT vs. HR-CONV with a willingness to pay threshold of 20,000 EUR (red diagonal line)

Panel b: ICUR per person comparing LR-CONV vs. LR-CTRL with a willingness to pay threshold of 20,000 EUR (red diagonal line)

Panel c: Probability of cost-effectiveness for high-risk and low-risk cohorts at different willingness to pay thresholds

References

1. Verband der Privaten Krankenversicherung (2013) Gebührenordnung für Ärzte (GOÄ) mit verkürzten Leistungsbezeichnungen. Available from<https://www.derprivatpatient.de/sites/default/files/gebuehrenordnung-fuer-aerzte.pdf>. Accessed 20 September 2023
2. Kassenärztliche Bundesvereinigung (2017) Einheitlicher Bewertungsmaßstab (EBM). Stand 3. Quartal 2017.Available from <https://www.kbv.de/media/sp/EBM_Gesamt_-_Stand_3._Quartal_2017.pdf>. Accessed 20 September 2023
3. Kassenärztliche Bundesvereinigung (2020) Abrechnungsstatistik der Kassenärztlichen Bundesvereinigung. Honorarbericht nach §87c SGB V 1. Quartal 2013 bis 2. Quartal 2019. Available from <https://www.kbv.de/html/honorarbericht.php>. Accessed 20 September 2023
4. Kassenzahnärztliche Bundesvereinigung (2018) Jahrbuch 2018.Statistische Basisdaten zur vertragszahnärztlichen Versorgung. Einschließlich GOZ-Analyse. https://www.kzbv.de/kzbv-jahrbuch-2018.media.21f2fd08cc7dbf0c07422eb110f022ea.pdf
5. Statistisches Bundesamt. H 1-Gesundheit: DRG-Statistik 2010-2018- Vollstationäre Patientinnen Und Patienten in Krankenhäusern. [Erlösvolumen-Nach Major Diagnostic Categories (MDCs)-Je Fall in Euro]; 2019.
6. Bundesministerium für Gesundheit (2003) Gesetz zur Modernisierung der gesetzlichen Krankenversicherung (GKV-Modernisierungsgesetz-GMG).Available from <https://www.bgbl.de/xaver/bgbl/start.xav?startbk=Bundesanzeiger_BGBl&start=//*%255B@attr_id=%2527bgbl103s2190.pdf%2527%255D#__bgbl__%2F%2F*%5B%40attr_id%3D%27bgbl103s2190.pdf%27%5D__1582107643968>. Accessed 20 September 2023
7. Bundesministerium für Gesundheit (2017) Ergebnisse der Statistik KG 5, Vorsorge- und Rehabilitationsmaßnahmen 2017 der Gesetzlichen Krankenversicherungen. Available from <https://www.bundesgesundheitsministerium.de/fileadmin/Dateien/3_Downloads/Statistiken/GKV/Geschaeftsergebnisse/Ergebnisse_der_Statistik_KG_5__Vorsorge-_und_Rehabilitationsmassnahmen_2017.pdf>. Accessed 20 September 2023
8. Bundesministerium für Gesundheit (2018)Gesetzliche Krankenversicherung.Endgültige Rechnungsergebnisse 2017. Available from <https://www.bundesgesundheitsministerium.de/fileadmin/Dateien/3_Downloads/Statistiken/GKV/Finanzergebnisse/KJ1_2017_Internet.pdf>. Accessed 20 September 2023 https://www.bundesgesundheitsministerium.de/themen/krankenversicherung/zahlen-und-fakten-zur-krankenversicherung/finanzergebnisse.html
9. Bundesministerium für Gesundheit (2023)Zuzahlungen der privaten Haushalte in der gesetzlichen Krankenversicherung. Available from <http://www.gbe-bund.de/gbe10/i?i=664:37524060D>. Accessed 20 September 2023
10. Wissenschaftliches Institut der AOK. GKV-Arzneimittelindex: Stammdatei Plus; 2018.
11. Statistisches Bundesamt (2018) Verdienste und Arbeitskosten. Arbeitnehmerverdienste 2017. Available from <https://www.statistischebibliothek.de/mir/receive/DEHeft_mods_00076453>. Accessed 20 September 2023
12. Deutsche Rentenversicherung Bund. Rentenversicherung in Zeitreihen.Oktober 2019. Available from <https://www.bmas.de/SharedDocs/Downloads/DE/Rente/Kommission-Verlaesslicher-Generationenvertrag/deutsche-rentenversicherung-rentenversicherung-in-zeitreihen.pdf;jsessionid=7B8150A166A09FF462E4D58D7A48C101.delivery1-master?__blob=publicationFile&v=1>. Accessed 20 September 2023
13. Statistisches Bundesamt. Verdienste und Arbeitskosten: Nettoverdienste-Modellrechnung-2017. https://www.destatis.de/DE/Themen/Arbeit/Verdienste/Realloehne-Nettoverdienste/Publikationen/Downloads-Realloehne-Nettoverdienste/nettoverdienste-modellrechnung-2160250187004.pdf?__blob=publicationFile
14. Bauer A, Fuchs J, Gartner H et al. (2021) IAB-Prognose 2021. Arbeitsmarkt auf dem Weg aus der Krise. IAB-Kurzbericht 06/2021. Available from <https://doku.iab.de/kurzber/2021/kb2021-06.pdf>. Accessed 20 September 2023
15. Seidl H, Bowles D, Bock J-O et al (2015) FIMA-questionnaire for health-related resource use in an elderly population: development and pilot study. 77(1):46-52. https://doi.org/ 10.1055/s-0034-1372618
16. Hernan WH, Brandle M, Zhang P, et al.(2003) Costs associated with the primary prevention of type 2 diabetes mellitus in the diabetes prevention program. Diabetes Care 26(1):36-47. https://doi.org/10.2337/diacare.26.1.36
17. Bock J-O, Brettschneider C, Seidl H et al. (2015) Calculation of standardised unit costs from a societal perspective for health economic evaluation. Gesundheitswesen 77(1):53-61. https://doi.org/10.1055/s-0034-1374621
18. Icks A, Chernyak N, Bestehorn K et al.(2010) Methods of Health Economic Evaluation for Health Services Research. Gesundheitswesen72(12):917-933. https://doi.org/ 10.1055/s-0030-1262859
19. Faria R, Gomes M, Epstein D, White IR (2014) A Guide to Handling Missing Data in Cost-Effectiveness Analysis Conducted Within Randomised Controlled Trials. PharmacoEconomics 32(12):1157-1170. https://doi.org/10.1007/s40273-014-0193-3
20. Raghunathan TE, Lepkowksi JM, Van Hoewyk J, Solenbeger P (2001) A multivariate technique for multiply imputing missing values using a sequence of regression models. Surv Methodo27(1):85-95
21. van Buuren S (2007)Multiple imputation of discrete and continuous data by fully conditional specification. Stat Methods Med Res. 16(3):219-242. https://doi.org/10.1177/096228020607446
22. Bouhlila DS, Sellaouti F(2013) Multiple imputation using chained equations for missing data in TIMSS: a case study. *Large-scale Assess Educ* 1(4):1-33. <https://doi.org/10.1186/2196-0739-1-4>
23. Hardt J, Herke M, Leonhart R (2012)Auxiliary variables in multiple imputation in regression with missing X: A warning against including too many in small sample research. BMC Med Res Methodol 12:184 https://doi.org/10.1186/1471-2288-12-184
24. MacNeil Vroomen J, Eekhout I, Dijkgraaf MG, et al (2016) Multiple imputation strategies for zero-inflated cost data in economic evaluations: Which method works best? Eur J Health Econ 17(8):939-950. https://doi.org/10.1007/s10198-015-0734-5
25. Royston P (2009) Multiple Imputation of Missing Values: Further Update of Ice, with an Emphasis on Categorical Variables. Stata J. 9(3):466-477. [https://doi.org/10.1177/1536867X0900900308](https://doi.org/10.1177/1536867X0900900308%20)
26. Duijzer G, Bukman AJ, Meints-Groenveld A et al.(2019) Cost-effectiveness of the SLIMMER diabetes prevention intervention in Dutch primary health care: economic evaluation from a randomised controlled trial. BMC Health Serv Res 19(1):824. <https://doi.org/10.1186/s12913-019-4529-8>
27. <https://www.gkv-spitzenverband.de/krankenversicherung/ambulante_leistungen/heilmittel/125_ernaehrung/125_ernaehrungstherapie.jsp>
